# Indirect impacts of the COVID-19 pandemic at two tertiary neonatal units in Zimbabwe and Malawi: an interrupted time series analysis

**DOI:** 10.1101/2021.01.06.21249322

**Authors:** Simbarashe Chimhuya, Samuel R. Neal, Gwendoline Chimhini, Hannah Gannon, Mario Cortina-Borja, Caroline Crehan, Deliwe Nkhoma, Tarisai Chiyaka, Emma Wilson, Tim Hull-Bailey, Felicity Fitzgerald, Msandeni Chiume, Michelle Heys

**Affiliations:** Child and Adolescent Health Unit, Faculty of Medicine and Health Sciences, University of Zimbabwe, Harare, Zimbabwe; Population, Policy and Practice Research and Teaching Department, UCL Great Ormond Street Institute of Child Health, University College London, UK; Parent and Child Health Initiative Trust, Lilongwe, Malawi; Biomedical Research and Training Institute, Harare, Zimbabwe; Infection, Immunity and Inflammation, UCL Great Ormond Street Institute of Child Health, University College London, UK; Department of Paediatrics, Kamuzu Central Hospital, Lilongwe, Malawi

**Keywords:** COVID-19, neonatal, NICU, low-income countries, developing countries, global health

## Abstract

**Background:** Deaths from COVID-19 have exceeded 1.8 million globally (January 2020). We examined trends in markers of neonatal care before and during the pandemic at two tertiary neonatal units in Zimbabwe and Malawi.

**Methods:** We analysed data collected prospectively via the NeoTree app at Sally Mugabe Central Hospital (SMCH), Zimbabwe, and Kamuzu Central Hospital (KCH), Malawi. Neonates admitted from 1 June 2019 to 25 September 2020 were included. We modelled the impact of the first cases of COVID-19 (Zimbabwe: 20 March 2020; Malawi: 3 April 2020) on number of admissions, gestational age and birth weight, source of admission referrals, prevalence of neonatal encephalopathy, and overall mortality.

**Findings:** The study included 3,450 neonates at SMCH and 3,350 neonates at KCH. Admission numbers at SMCH did not initially change after the first case of COVID-19 but fell by 48% during a nurses’ strike (Relative risk (RR) 0·52, 95%CI 0·40-0·68, *p* < 0·002). At KCH, admissions dropped by 42% (RR 0·58; 95%CI 0·48-0·70; *p* < 0·001) soon after the first case of COVID-19. At KCH, gestational age and birth weight decreased slightly (1 week, 300 grams), outside referrals dropped by 28%, and there was a slight weekly increase in mortality. No changes in these outcomes were found at SMCH.

**Interpretation:** The indirect impacts of COVID-19 are context-specific. While this study provides vital evidence to inform health providers and policy makers, national data are required to ascertain the true impacts of the pandemic on newborn health.

**Funding:** International Child Health Group, Wellcome Trust.

**RESEARCH IN CONTEXT:** *Evidence before this study:* We searched PubMed for evidence of the indirect impact of the COVID-19 pandemic on neonatal care in low-income settings using the search terms *neonat\** **or** *newborn*, **and** *COVID-19* **or** *SARS-CoV 2* **or** *coronavirus*, and the Cochrane low and middle income country (LMIC) filters, with no language limits between 01.10.2019 and 21.11.20. While there has been a decrease in global neonatal mortality rates, the smaller improvements seen in low-income settings are threatened by the direct and indirect impact of the COVID-19 pandemic. A modelling study of this threat predicted between 250000-1.1 million extra neonatal deaths as a result of decreased service provision and access in LMICs. A webinar and survey of frontline maternal/newborn healthcare workers in >60 countries reported a decline in both service attendance and in quality of service across the ante-, peri- and post-natal journey. Reporting fear of attending services, and difficulty in access, and a decrease in service quality due to exacerbation of existing service weaknesses, confusion over guidelines and understaffing. Similar findings were reported in a survey of healthcare workers providing childhood and maternal vaccines in LMICs. One study to date has reported data from Nepal describing an increase in stillbirths and neonatal deaths, with institutional deliveries nearly halved during lockdown.

*Added value of this study:* To our knowledge, this is the first and only study in Sub-Saharan Africa describing the impact of COVID-19 pandemic on health service access and outcomes for newborns in two countries. We analysed data from the digital quality improvement and data collection tool, the NeoTree, to carry out an interrupted time series analysis of newborn admission rates, gestational age, birth weight, diagnosis of hypoxic ischaemic encephalopathy and mortality from two large hospitals in Malawi and Zimbabwe (*n*∼7000 babies). We found that the indirect impacts of COVID-19 were context-specific. In Sally Mugabe Central Hospital, Zimbabwe, initial resilience was demonstrated in that there was no evidence of change in mortality, birth weight or gestational age. In comparison, at Kamuzu Central Hospital, Malawi, soon after the first case of COVID-19, the data revealed a fall in admissions (by 42%), gestational age (1 week), birth weight (300 grams), and outside referrals (by 28%), and there was a slight weekly increase in mortality (2%). In the Zimbabwean hospital, admission numbers did not initially change after the first case of COVID-19 but fell by 48% during a nurses’ strike, which in itself was in response to challenges exacerbated by the pandemic.

*Implications of all the available evidence:* Our data confirms the reports from frontline healthcare workers of a perceived decline in neonatal service access and provision in LMICs. Digital routine healthcare data capture enabled rapid profiling of indirect impacts of COVID-19 on newborn care and outcomes in two tertiary referral hospitals, Malawi and Zimbabwe. While a decrease in service access was seen in both countries, the impacts on care provided and outcome differed by national context. Health systems strengthening, for example digital data capture, may assist in planning context-specific mitigation efforts.

## INTRODUCTION

The World Health Organization declared coronavirus disease (COVID-19) a Public Health Emergency of International Concern on 30 January 2020.^1^ Confirmed cases have exceeded 80 million globally with nearly two million deaths.^2^ Zimbabwe recorded its first case on 20 March and has reported >17000 cases with >400 deaths to date.^2^ Malawi confirmed its first three cases on 3 April and has reported ∼7000 cases and ∼200 deaths to date.^2^

Before the COVID-19 pandemic, considerable improvements were made in global child health: the global neonatal mortality rate fell from 31 to 18 deaths per 1,000 live births between 2000 and 2018.^3^ Yet there were disparities in the rates of decline with the sub-Saharan Africa region facing highest neonatal mortality rates^3^. Now, there is a danger that health outcomes in low-income and middle-income countries (LMICs) will fall further behind high-income countries. While countries worldwide face challenges related to the COVID-19 pandemic, LMICs are particularly struggling with financial constraints, limited testing capacity, lack of personal protective equipment, and staff shortages.^4,5^ As children are at low-risk of infection or severe disease from COVID-19,^6-10^ any impacts on their health outcomes will likely be attributable to the indirect effects of the pandemic on health systems, as in previous disease outbreaks.^11,12^ These include increased rates of parental unemployment, food and housing insecurity, and reduced access to routine care.^13,14^

The NeoTree application (app) is an Android tablet-based quality improvement platform that aims to reduce neonatal mortality in LMICs.^15^ Developed in collaboration with local stakeholders, it is embedded in routine practice at two neonatal units (NNUs) in Zimbabwe and Malawi, providing real-time clinical decision support, neonatal care education, and digital data capture.^16,17^

We aimed to examine trends in markers of neonatal care before and during the COVID-19 pandemic at Sally Mugabe Central Hospital (SMCH), Zimbabwe, and Kamuzu Central Hospital (KCH), Malawi. Specifically, we compared the:

1. number of admissions,
2. gestational age and birth weight of admitted neonates,
3. source of admission referrals,
4. prevalence of neonatal encephalopathy (NE), and
5. overall mortality rate

before and after the first reported cases of COVID-19.

## METHODS

This study is reported in accordance with the Strengthening the Reporting of Observational Studies in Epidemiology (STROBE) statement (Appendix 1).

### Setting

SMCH is a public referral hospital in Harare, Zimbabwe. It has the largest of three tertiary NNUs nationwide with 100 cots and predominantly doctor-led care. KCH, Lilongwe, is one of four regional referral hospitals in Malawi and the NNU has 75 cots. In contrast to SMCH, care in the NNU is mostly nurse-led. Both units accept local and national referrals for specialist surgical care.

### Participants

All neonates admitted to each NNU over a 16-month period from 1 June 2019 to 25 September 2020 (69 complete weeks) were eligible for inclusion. We applied no specific exclusion criteria.

### Data collection

Data were collected prospectively using the NeoTree app. Health workers complete a digital form when a neonate is admitted to the unit (admission form) and when they are discharged or die (outcome form). The app guides assessment of the neonate and collects data on patient demographics, examination findings, diagnoses, and interventions. Pseudonymised forms are uploaded monthly to University College London servers (Zimbabwe data) and Amazon Web Services (Malawi data). Admission and outcome forms are linked by a unique identifier generated by the app at admission.

### Outcomes

We evaluated five outcomes:

1. Number of admissions: determined from the admission date of each completed admission form.
2. Gestational age at birth (weeks) and birth weight (grams): as entered into the admission form from obstetric records.
3. Source of admission: defined as ‘within’ (labour ward, postnatal ward, antenatal ward, obstetric theatre, or fee-paying ward [KCH only]) or ‘outside’ (referral from another health facility or postnatal self-referral from home).
4. Diagnosis of NE: defined as “hypoxic ischaemic encephalopathy” or “birth asphyxia” recorded as a diagnosis, cause of death or contributory cause of death on the outcome form.
5. Mortality: defined as an outcome of “neonatal death” on the outcome form. All other neonates, including those discharged, transferred to another facility or who left on parental request, were considered alive.

### Ethical approval

Research ethics approval was granted by the UCL Research Ethics Committee (17123/001) and ethics committees in Malawi (P.01/20/2909) and Zimbabwe (MRCZ/A/2570) (Appendix 2). The need to obtain informed consent was waived as we collected only pseudonymised data routinely documented for clinical care.

### Statistical analysis

Analyses were performed in R version 3.6.3,^18^ running on RStudio version 1.2.5033.^19^ First, admission forms were matched with their corresponding outcome form based on the unique identifier generated at admission. Lack of completed outcome forms (SMCH: *n*=316[9.1%]; KCH: *n*=243[7.2%]) or errors in entry of the unique identifier at discharge (SMCH: *n*=318[9.2%]; KCH: *n*=182[5.4%]) meant we were unable to match some admission forms with outcome forms (SMCH: *n*=634[18.3%]; KCH: *n*=425[12.6%]). For outcomes 1-3, we based analyses on data from all admission forms, regardless of match status. For outcomes 4 and 5, we based analyses on matched records only. Matched records implying a negative admission duration (i.e. outcome date prior to admission date) were excluded (SMCH: *n*=58[2%]; KCH: *n*=25[1%]). See Appendix 3 for a flow diagram of record inclusion. Missing data were excluded using pairwise deletion for each analysis as frequencies of missing values were minimal (Appendix 4).

This study used an interrupted time series design with weekly data windows. We considered the first confirmed case of COVID-19 in each country as the intervention (Zimbabwe: 20 March 2020; Malawi: 3 April 2020).^2^ For all outcomes, we hypothesised a level change impact model without a lag (for a description of these models, see Bernal et al.^20^). Gestational age and birth weight were modelled with linear regression. All other outcomes were modelled using quasi-Poisson regression to account for overdispersion,^21^ with the logarithm of the number of admissions in each weekly window included as an offset. All SMCH models were adjusted for a period of doctors’ strikes from 3 September 2019 to 22 January 2020.^22^ KCH models were unadjusted. Additional models were constructed to explore the effects of a nurses’ strike in Zimbabwe (17 June to 9 September 2020)^23^ and alternative impact models. Nested models were compared with the *F*-test. See Appendix 5 for model details.

## RESULTS

### Outcome 1: Admissions to the neonatal unit

We included 3,450 neonates at SMCH and 3,350 neonates at KCH. Figure 1 shows the seven-day moving average of admissions to the NNU.

**Figure 1:**
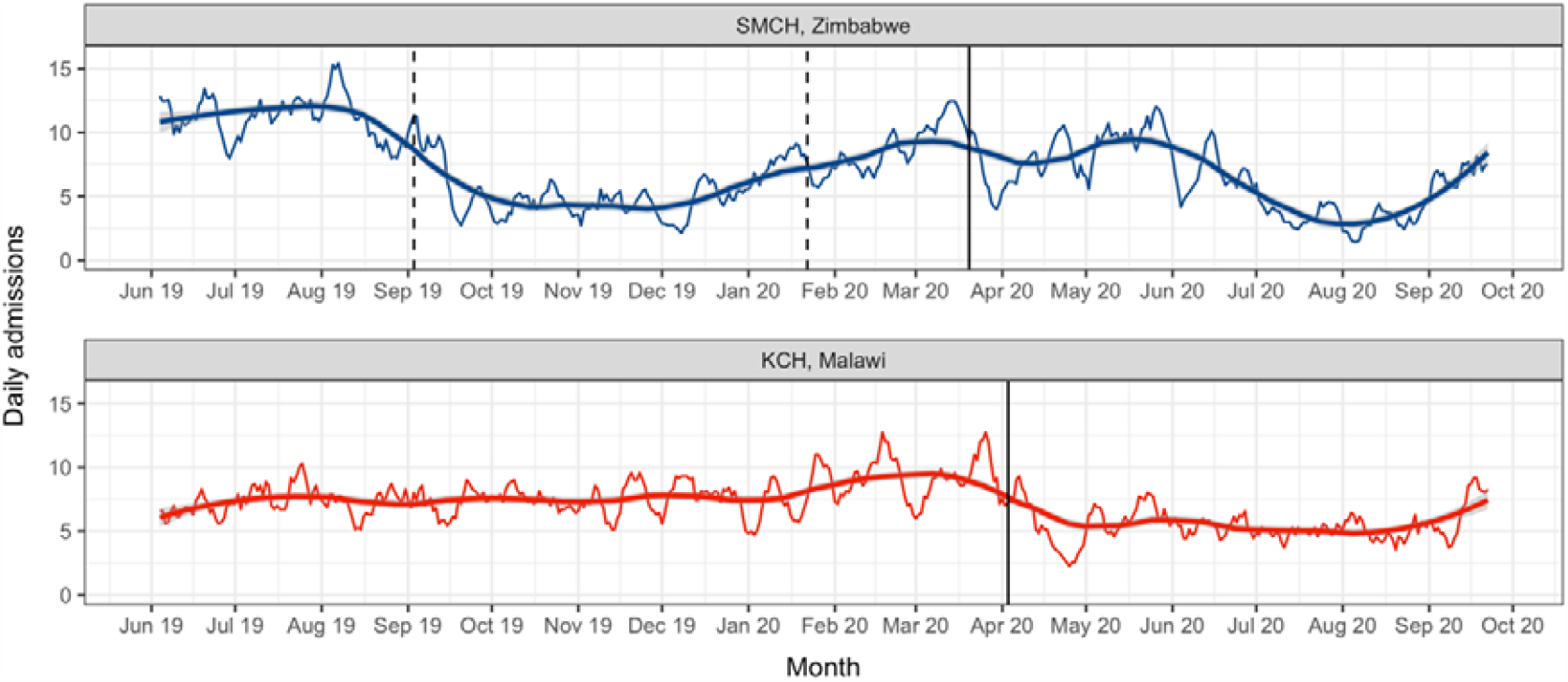
Trend in daily admissions to the neonatal unit. • The seven-day moving average of daily admission numbers has been plotted. • Smoothed line: local regression (LOESS) model fitted on the seven-day moving average of daily admission numbers; shaded region: 95% confidence interval. • Solid vertical line: first confirmed case of COVID-19 in each country. • Period between dashed vertical lines: industrial action by doctors in Zimbabwe. • Counts based on all admission forms completed, irrespective of match status. • *SMCH: Sally Mugabe Central Hospital; KCH: Kamuzu Central Hospital*

At SMCH, the mean (SD) number of weekly admissions was 54·6 (23·5) before the first case of COVID-19 (pre-COVID-19) and 42·8 (19·9) afterwards (post-COVID-19). The level change regression model, adjusted for the doctors’ strike, showed no evidence of a change in admissions after the first case of COVID-19 (relative risk [RR] 0·83; 95% confidence interval [CI] 0·60-1·14; *p* = 0·25) but the scatterplot indicated this model fit the data poorly (model 1, Figure 2A). An alternative model, additionally adjusted for the nurses’ strike, again showed no change in the overall post-COVID-19 period (RR 0·90; 95%CI 0·69-1·17; *p* = 0·43) (model 2, Figure 2B). However, this model suggested that admissions fell by 48% during the nurses’ strike period (RR 0·52, 95%CI 0·40-0·68, *p* < 0·001) and fit the data better (*F*[1, 64] = 24·66, *p* < 0·001).

**Figure 2:**
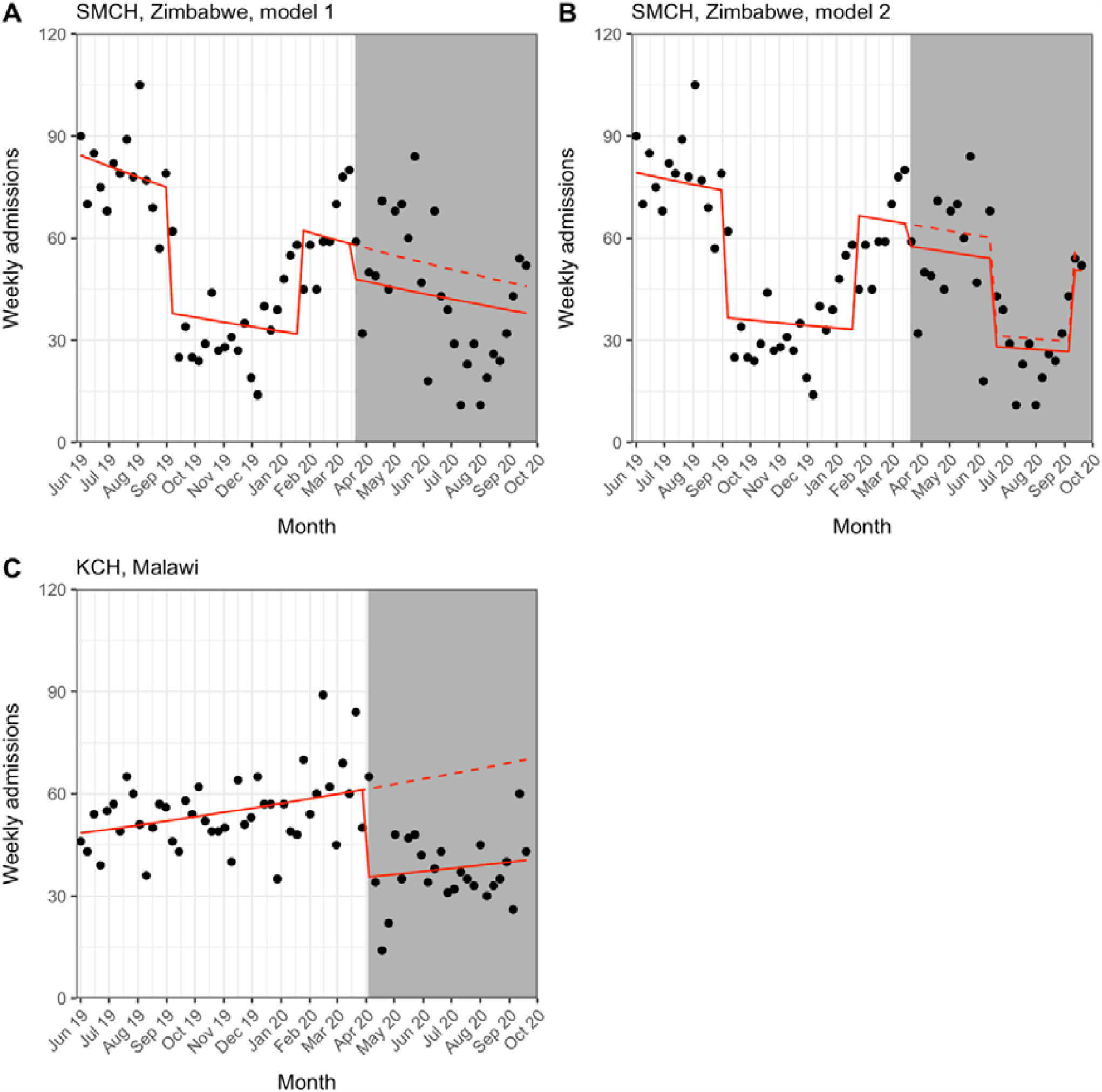
Interrupted time series for weekly admissions to the neonatal unit. • White background: pre-COVID-19 period; grey background: post-COVID-19 period. • Solid line: predicted trend from regression model; dashed line: counterfactual scenario. • SMCH model 1 (panel A) adjusted for doctors’ strike period; SMCH model 2 (panel B) additionally adjusted for nurses’ strike period; KCH model (panel C) unadjusted. • Counts based on all admission forms completed, irrespective of match status. • *SMCH: Sally Mugabe Central Hospital; KCH: Kamuzu Central Hospital*

At KCH, the mean (SD) number of weekly admissions was 54·5 (10·8) in the pre-COVID-19 period and 38·0 (10·9) in the post-COVID-19 period. The level change model suggested a 42% reduction in admissions after the first case of COVID-19 (RR 0·58; 95%CI 0·48-0·70; *p* < 0·001) (Figure 2C).

### Outcome 2: Gestational age and birth weight

At SMCH, the mean (SD) gestational age at birth was 36·1 (4·4) weeks in the pre-COVID-19 period and 36·0 (4·2) weeks in the post-COVID-19 period. The mean (SD) birth weight was 2500 (908) grams in the pre-COVID-19 period and 2487 (896) grams in the post-COVID-19 period. Regression analysis indicated no change in gestational age at birth nor birth weight after the first case of COVID-19 (gestational age: beta 0·04; 95%CI −0·53-0·61; *p* = 0·89, birth weight: beta −7·2; 95%CI −127·1-112·6; *p* = 0·91) (Figure 3A, Figure 3C). Adjusting for the nurses’ strike did not improve model fit (data not shown).

**Figure 3:**
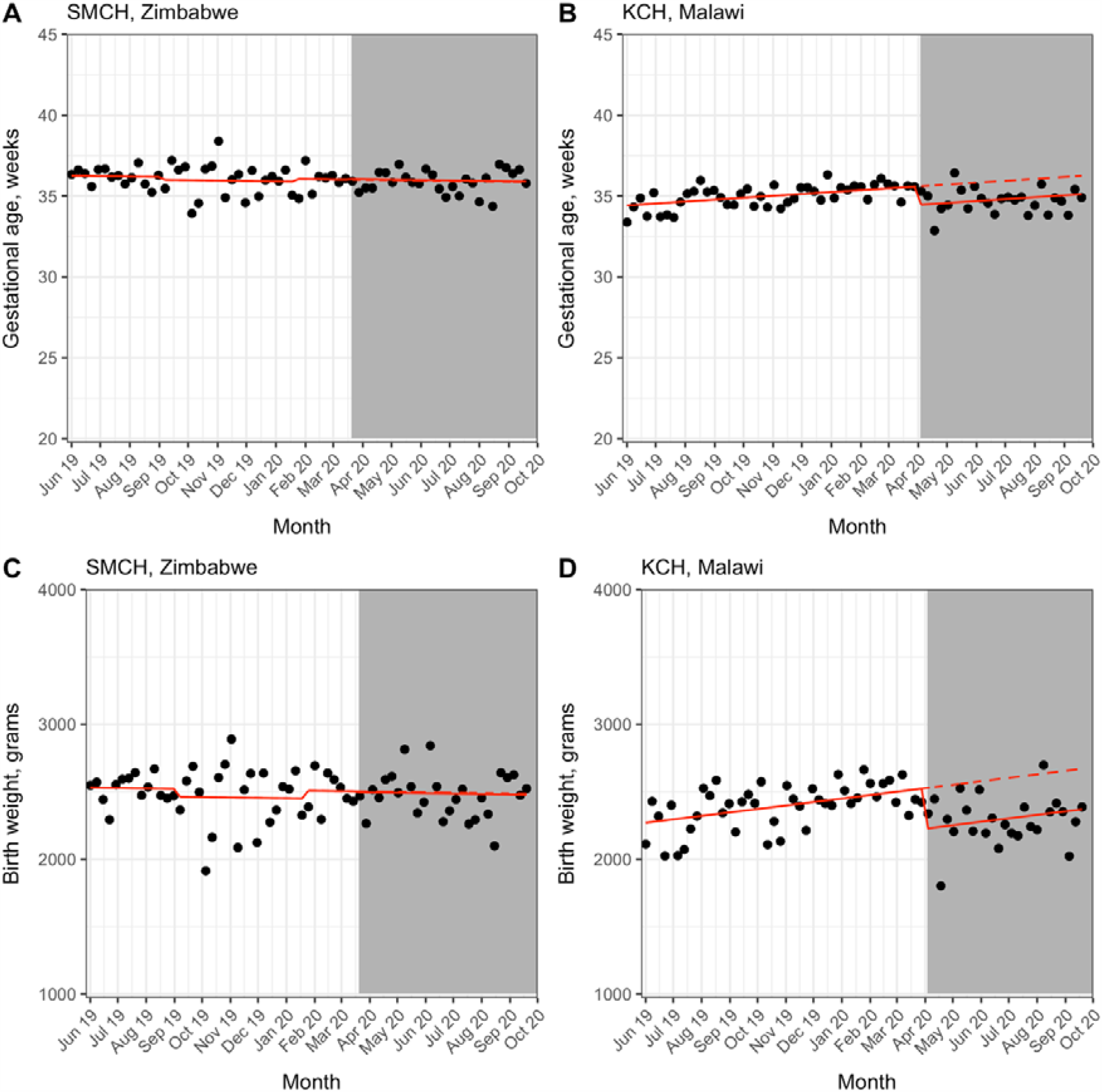
Interrupted time series for gestational age and birth weight. • Data points represent weekly mean gestational age or birth weight to avoid overplotting. • White background: pre-COVID-19 period; grey background: post-COVID-19 period. • Solid line: predicted trend from regression model; dashed line: counterfactual scenario. • SMCH models (panels A & C) adjusted for doctors’ strike period, KCH models (panels B & D) unadjusted. • Data from all admission forms completed, irrespective of match status. • *SMCH: Sally Mugabe Central Hospital; KCH: Kamuzu Central Hospital*

At KCH, the mean (SD) gestational age was 35·0 (3·9) weeks in the pre-COVID-19 period and 34·8 (3·9) weeks in the post-COVID-19 period. The mean (SD) birth weight was 2402 (883) grams in the pre-COVID-19 period and 2299 (870) grams in the post-COVID-19 period. Gestational age decreased by one week in the post-COVID-19 period (beta −1·14; 95%CI −1·62-[-]0·65; *p* < 0·001) (Figure 3B) and birth weight decreased by 300 grams (beta −299·9; 95%CI −412·3-[-]187·5; *p* < 0·001) (Figure 3D).

### Outcome 3: Source of admission referral

At SMCH, the mean (SD) percentage of outside referrals to the NNU was 39(11)% in the pre-COVID-19 period and 35(9)% in the post-COVID-19 period. The regression model showed no evidence of a change in the percentage of outside referrals after the first case of COVID-19 (RR 0·98; 95%CI 0·79-1·23; *p* = 0·88) (Figure 4A). Adjusting for the nurses’ strike did not improve model fit (data not shown).

**Figure 4:**
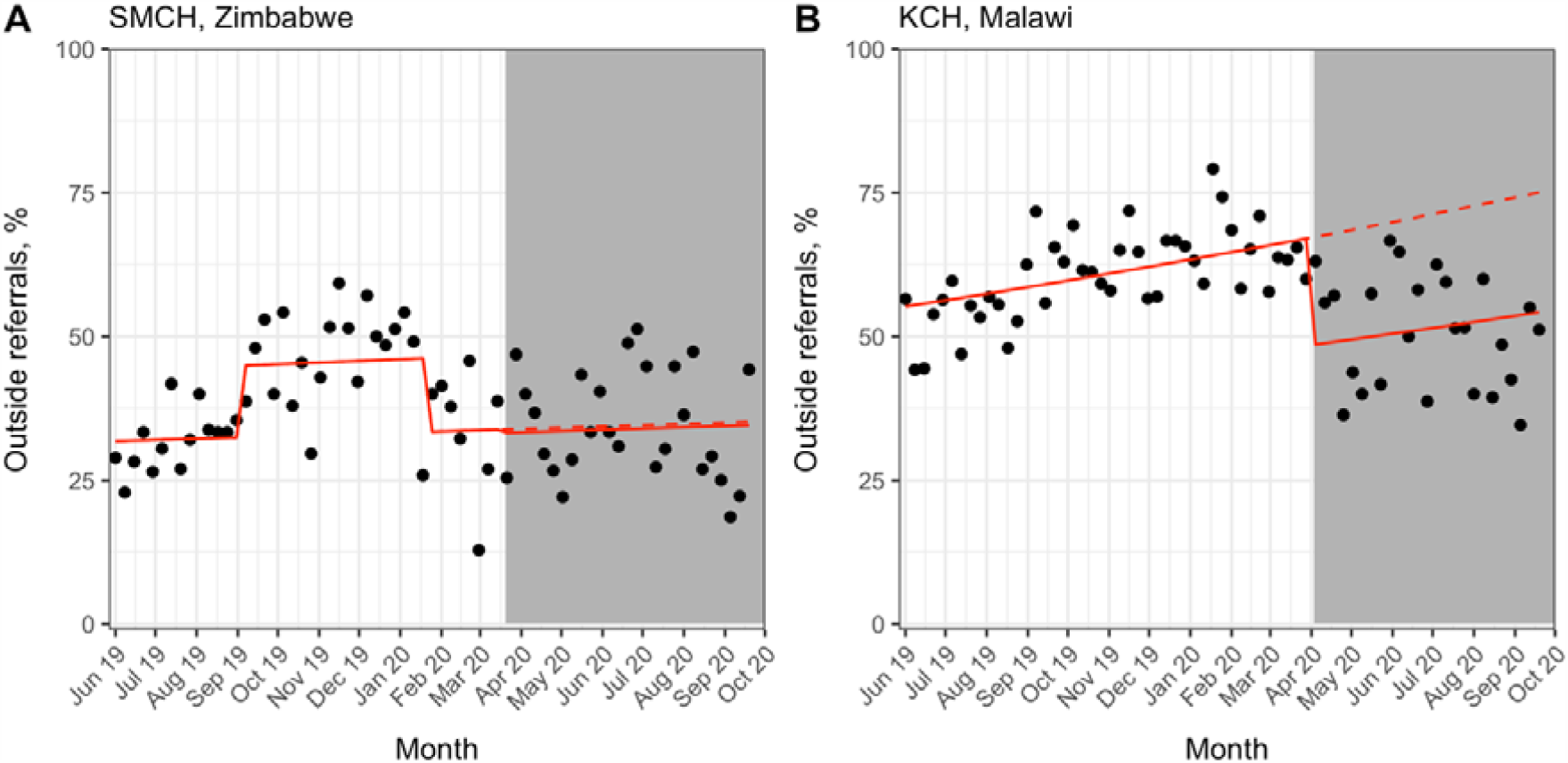
Interrupted time series for outside referrals to the neonatal unit. • White background: pre-COVID-19 period; grey background: post-COVID-19 period. • Solid line: predicted trend from regression model; dashed line: counterfactual scenario. • SMCH model (panel A) adjusted for doctors’ strike period, KCH model (panel B) unadjusted. • Data from all admission forms completed, irrespective of match status. • *SMCH: Sally Mugabe Central Hospital; KCH: Kamuzu Central Hospital*

At KCH, the mean (SD) percentage of outside referrals was 61(8)% in the pre-COVID-19 period and 51(10)% in the post-COVID-19 period. Regression analysis suggested a 28% relative reduction in outside referrals after the first case of COVID-19 (RR 0·72; 95%CI 0·65-0·81; *p* < 0·001) (Figure 4B).

### Outcome 4: Prevalence of neonatal encephalopathy

At SMCH, the mean (SD) percentage of admitted neonates diagnosed with NE was 16(6)% in the pre-COVID-19 period and 21(12)% in the post-COVID-19 period suggesting a possible increase. Regression analysis showed no statistically significant change in the percentage of neonates diagnosed with NE post-COVID-19 (RR 1·08; 95%CI 0·76-1·55; *p* = 0·67) (Figure 5A). Adjusting for the nurses’ strike did not improve model fit (data not shown).

**Figure 5:**
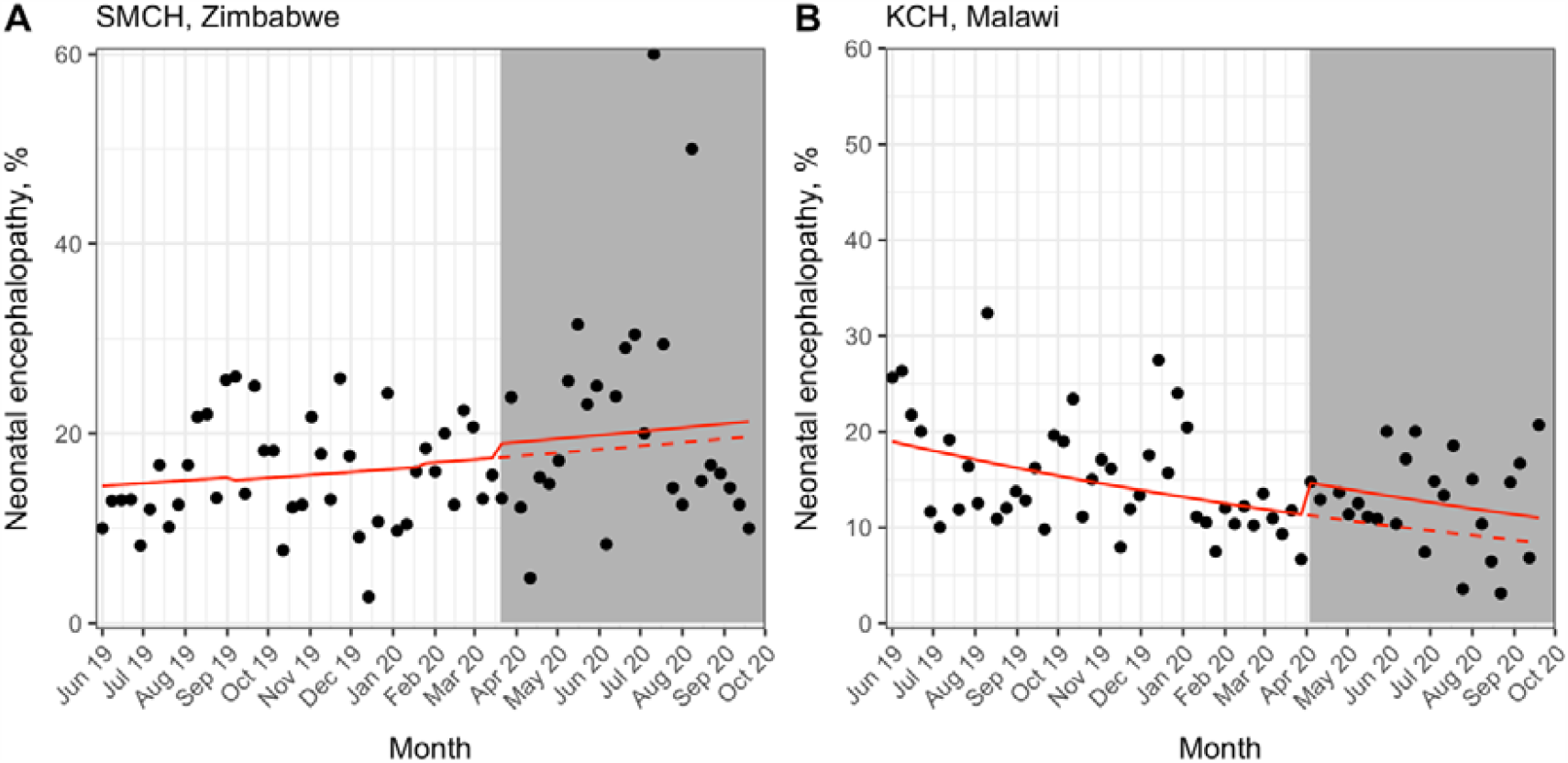
Interrupted time series for prevalence of neonatal encephalopathy. • White background: pre-COVID-19 period; grey background: post-COVID-19 period. • Solid line: predicted trend from regression model; dashed line: counterfactual scenario. • SMCH model (panel A) adjusted for doctors’ strike period, KCH model (panel B) unadjusted. • Data from matched admission and outcome forms only. • *SMCH: Sally Mugabe Central Hospital; KCH: Kamuzu Central Hospital*

At KCH, the mean (SD) percentage of admitted neonates diagnosed with NE was 15(6)% in the pre-COVID-19 period and 13(5)% in the post-COVID-19 period. The regression model suggested a possible increase in diagnoses of NE after the first case of COVID-19, but this was not statistically significant (RR 1·30; 95%CI 0·95-1·80; *p* = 0·11) (Figure 5B).

### Outcome 5: Overall mortality

For SMCH, the mean (SD) percentage of deaths per week of admission was 25(10)% in the pre-COVID-19 period and 26(16)% in the post-COVID-19 period. The level change regression model, adjusted for the doctors’ strike, showed no evidence of a change in mortality after the first case of COVID-19 (RR 0·80; 95%CI 0·56-1·15; *p* = 0·23) but the scatterplot indicated this model fit the data poorly (model 1, Figure 6A). An alternative model, additionally adjusted for the nurses’ strike, again showed no change in overall mortality (RR 0·72; 95%CI 0·51-1·03; *p* = 0·07) but fit the data better (F[1, 64] = 11·61, *p* = 0·001) (model 2, Figure 6B).

**Figure 6:**
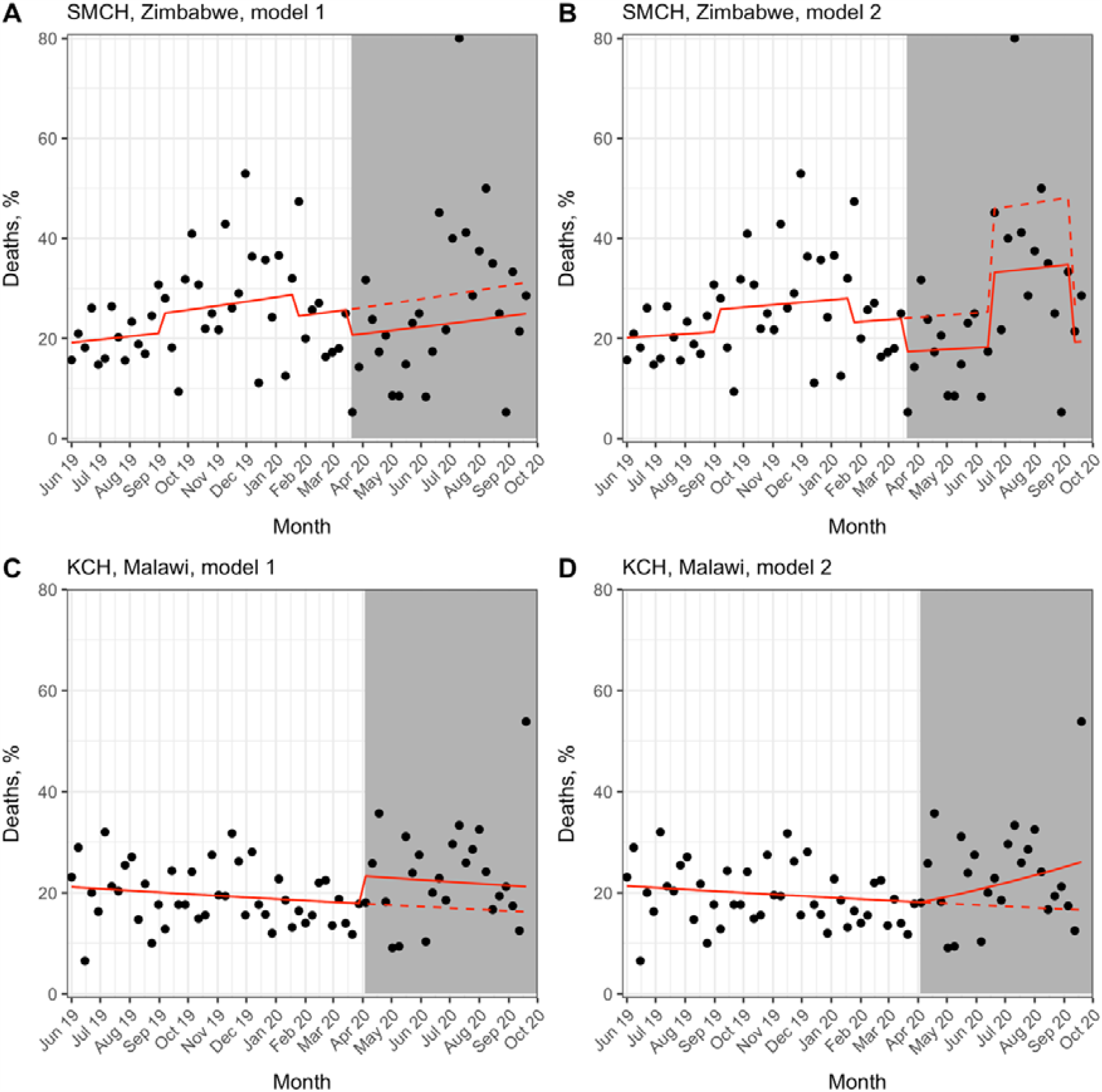
Interrupted time series for overall mortality. • White background: pre-COVID-19 period; grey background: post-COVID-19 period. • Solid line: predicted trend from regression model; dashed line: counterfactual scenario. • SMCH model 1 (panel A) adjusted for doctors’ strike period; SMCH model 2 (panel B) additionally adjusted for nurses’ strike period; KCH model 1 (panel C) unadjusted level change model; KCH model 2 (panel D) unadjusted slope change model. • Data from matched admission and outcome forms only. • *SMCH: Sally Mugabe Central Hospital; KCH: Kamuzu Central Hospital*

For KCH, the mean (SD) percentage of deaths per week of admission was 19(6)% in the pre-COVID-19 period and 23(9)% in the post-COVID-19 period. The level change regression model suggested a possible increase in mortality after the first case of COVID-19, but this was not statistically significant (RR 1·31; 95%CI 0·98-1·73; *p* = 0·07) (Figure 6C). However, fitting a slope change impact model suggested a small relative increase in mortality by 2% per week in the post-COVID-19 period (RR 1·02 per week; 95%CI 1·00-1·04, *p* = 0·04) (Figure 6D).

## DISCUSSION

### Summary

We performed an interrupted time series analysis to examine changes in neonatal care provision at two tertiary NNUs in Zimbabwe and Malawi after the first cases of COVID-19. We found that admissions at SMCH did not change significantly after the first case of COVID-19 when considering this period as a whole, but there was a considerable decrease (∼50%) in the number admissions in June to August 2020, coinciding with a nurses’ strike. We did not find significant changes in gestational age or birth weight, source of admission referrals, prevalence of NE or mortality at SMCH. Conversely, we found several changes in markers of neonatal care at KCH after the first case of COVID-19 in Malawi. The number of admissions fell by 42% and we noted a decrease in the gestational age and birth weight of admitted neonates (by ∼1 week and ∼300 grams, respectively), a 28% relative decrease in outside referrals, and a small but statistically significant weekly increase in mortality by 2% after the first case of COVID-19. Although this study is descriptive, we can speculate about explanations for our results based on existing literature and discussions with local health workers.

### Interpretation

The number of admissions at SMCH fell by around 50% between June to August 2020, but we noted no change outside this strike period, suggesting some resilience to the impact of the pandemic. However, nurses went on strike over pay and availability of personal protective equipment,^23^ so the strike is itself an indirect consequence of COVID-19. A similar reduction in admissions was seen at KCH, but, unlike at SMCH, this 42% decrease was noted within a week of the first case of COVID-19. In Figure 7, we propose several interlinked factors that might explain reduced admissions to the NNU. Several of these factors, such as fear of using health services, disrupted transport networks and staff shortages have been directly reported by local sources in LMICs and were highlighted in a recent report by Graham et al.^24^

**Figure 7:**
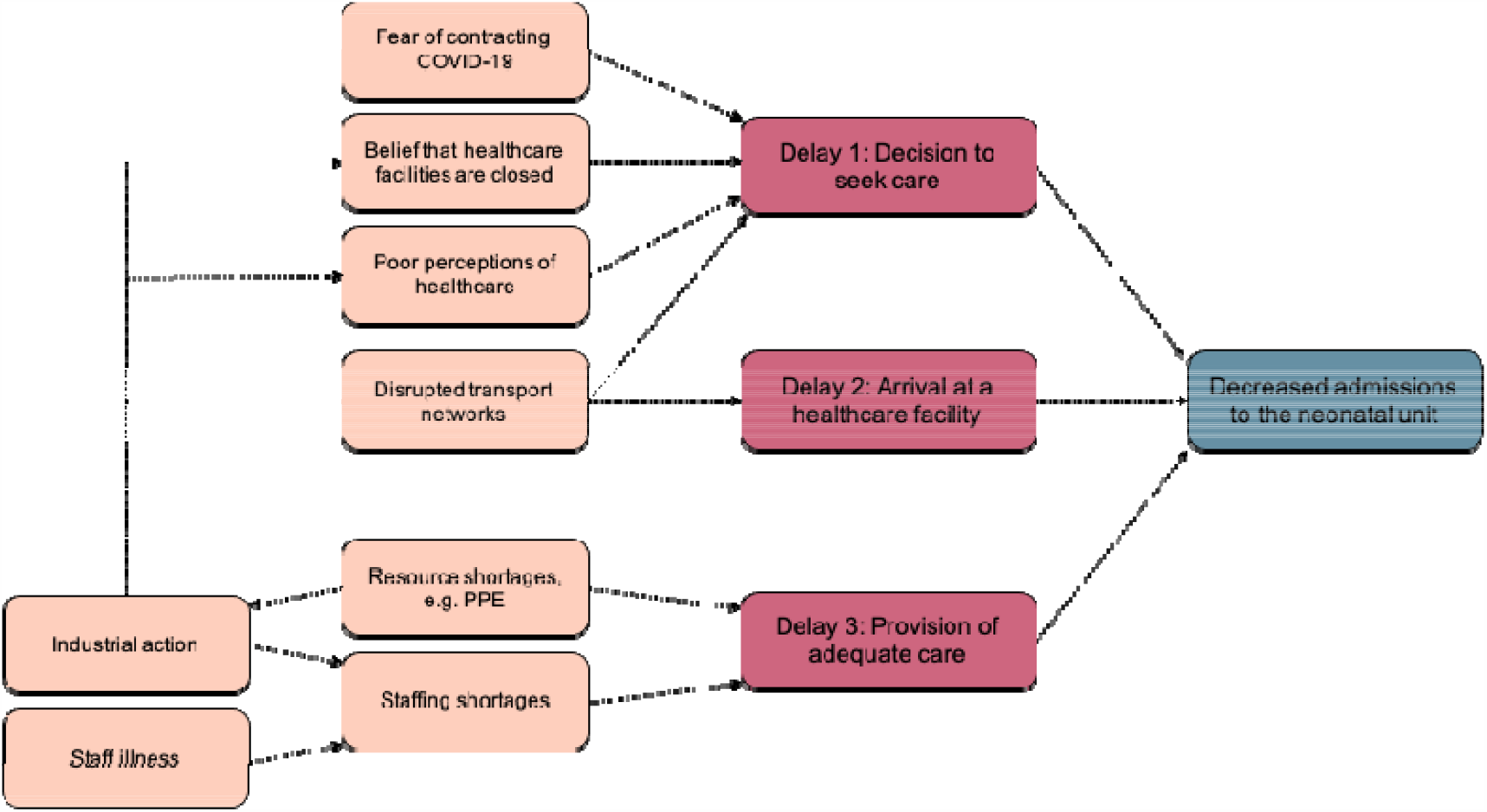
Possible factors influencing the decrease in admissions to the neonatal unit. • Delays (red boxes) derived from the “Three Delays” model of pregnancy-related mortality.^25^ • *COVID-19: coronavirus disease 2019; PPE: personal protective equipment*

We found a slight decrease in gestational age and birth weight of neonates at KCH, but not SMCH. Studies have reported increased rates of preterm birth in pregnant women with COVID-19 compared to those without the disease, mostly from medically-induced preterm birth; although none of these studies were conducted in LMICs.^26^ Preliminary analysis suggests rates of emergency caesarean section increased at SMCH and KCH, with a more marked increase at KCH (Appendix 6).

This is one potential explanation for our findings. However, we noted that the number of outside referrals decreased by 28% at KCH, and neonates referred from outside KCH are more likely to be from lower-risk pregnancies that delivered in a health centre with higher gestational ages and birth weights. Further analysis should stratify by source of admission referral to clarify this finding, but is supported by the fact that referrals were rigorously triaged by the on-call paediatrician during the pandemic, and that referrals from some areas were diverted away from KCH.

We hypothesised that rates of NE would increase during the pandemic. NE is the clinical manifestation of disordered brain function and can have multiple aetiologies.^27^ The term ‘hypoxic-ischaemic encephalopathy’ is reserved for cases where there is evidence of intrapartum asphyxia.^27^ In LMICs, obstructed labour is a major cause of maternal mortality and can lead to intrapartum asphyxia with subsequent neonatal morbidity and mortality, including NE.^28^ Therefore, the prevalence of NE might be expected to increase as a marker of delayed presentation to a health facility. It is reassuring that we did not find increased rates of NE at SMCH or KCH. However, these findings should be interpreted cautiously as some neonates with NE may not have presented to a health facility at all.

Finally, we observed a slight increase in overall mortality at KCH (a relative increase of 2% per week after the first case of COVID-19), although not at SMCH. In KCH, the increase in mortality may be due to decreased gestational age and birthweight, but also due to a reduced rota of nursing staff implemented to protect healthcare workers. In fact, there was a suggestion that mortality decreased after the first case of COVID-19 in Zimbabwe, but this was not statistically significant. The reasons for this are unclear but could include factors such as increased stillbirth rates or improved care for the smaller number of neonates on the NNU. More complete analysis of facility-based and community-based neonatal mortality is greatly needed.

### Limitations and future work

Some limitations should be noted. A limitation intrinsic to interrupted time series analysis is the possibility that another event occurred close to the first case of COVID-19 in either country causing spurious observations. Another potential threat to validity is changing data collection practices. For example, overstretched clinicians might not input data into the NeoTree app for all admitted neonates. However, this is unlikely as the NeoTree app is embedded into routine practice at SMCH and KCH and discussions with local collaborators suggest use of the app has continued without issue.

The NeoTree app only collects data on neonates admitted to the NNU. Therefore, our analysis does not capture stillbirths or neonatal deaths that occur in the community. It is troubling to see a dramatic fall in admissions in both sites, raising the possibility that many unwell neonates did not attend a health facility and died at home. A recent study found that facility births decreased by over 50% during the lockdown in Nepal, and facility stillbirth and neonatal mortality rates increased significantly.^29^ The NeoTree research team is currently collecting data on stillbirths at SMCH and KCH, but these data will still only represent stillbirths that occurred in a health facility. Given the COVID-19 pandemic is not over, it will be important to repeat our analysis over the coming months to further examine longer-term trends in neonatal care provision.

## Conclusion

The indirect impacts of COVID-19 are context-specific, with more significant and evident effects on neonatal care provision seen at KCH (Malawi) than SMCH (Zimbabwe). While this study provides vital evidence to inform health providers and policy makers, national data are required to ascertain the true impacts of the pandemic on newborn health.

## Data Availability

Data sharing statement
Data collected for the study cannot yet be made publicly available yet because primary analysis for the pilot implementation evaluation of the NeoTree, as well as secondary analysis are ongoing. A goal of our pilot implementation is the establishment of an open-source anonymised research database of data collected using the NeoTree in order to maximise the reach and utility for researchers aiming to improve outcomes for neonates in low income settings. This database is under development and subject to negotiation with relevant Ministries of Health.

## LIST OF ABBREVIATIONS

app: application
CI: confidence interval
COVID-19: coronavirus disease 2019
KCH: Kamuzu Central Hospital
LMIC: low-income and middle-income country
NE: neonatal encephalopathy
NNU: neonatal unit
RR: Relative risk
SD: standard deviation
SMCH: Sally Mugabe Central Hospital

## Contributors’ statement

Concept and study design by SC, SRN, GC, FF, MCB, CC, MC and MH with input from other authors. Data collected by HG, DN, TC, CC and THB. Analysis performed by SRN and MCB with contributions from FF, SC, EW & MH. Manuscript drafted by SC and SRN with input from GC, FF, MCB, MC & MH. All authors proof-read and approved final draft. Underlying data accessed and verified by SRN, MCB, HG, FF & MH.

## Acknowledgements

We would like to thank the funders of this study. Mr S. R. Neal was awarded the International Child Health Group David Morley Elective Bursary for this elective project. Funders of the wider NeoTree project, past and present, include the Wellcome Trust Digital Innovation Award (215742/Z/19/Z: PI: Heys), RCPCH, Naughton-Cliffe Mathews, UCL Grand Challenges and Global Engagement Fund, and the Healthcare Infection Society (SRG 201802004). Dr F. Fitzgerald is supported by the Academy of Medical Sciences and the funders of the Starter Grants for Clinical Lecturers scheme. This study and Drs M. Heys and F. Fitzgerald are further supported by the National Institute for Health Research Great Ormond Street Hospital Biomedical Research Centre. The funders had no role in study design, data collection and analysis, or preparation of this report. We are very grateful to the families at SMCH and KCH, and the staff members at both hospitals for their enthusiasm and commitment to the NeoTree project, without which this work would not be possible.

## Data sharing statement

Data collected for the study cannot yet be made publicly available yet because primary analysis for the pilot implementation evaluation of the NeoTree, as well as secondary analysis are ongoing. A goal of our pilot implementation is the establishment of an open-source anonymised research database of data collected using the NeoTree in order to maximise the reach and utility for researchers aiming to improve outcomes for neonates in low income settings. This database is under development and subject to negotiation with relevant Ministries of Health.

## APPENDIX 1: STROBE CHECKLIST

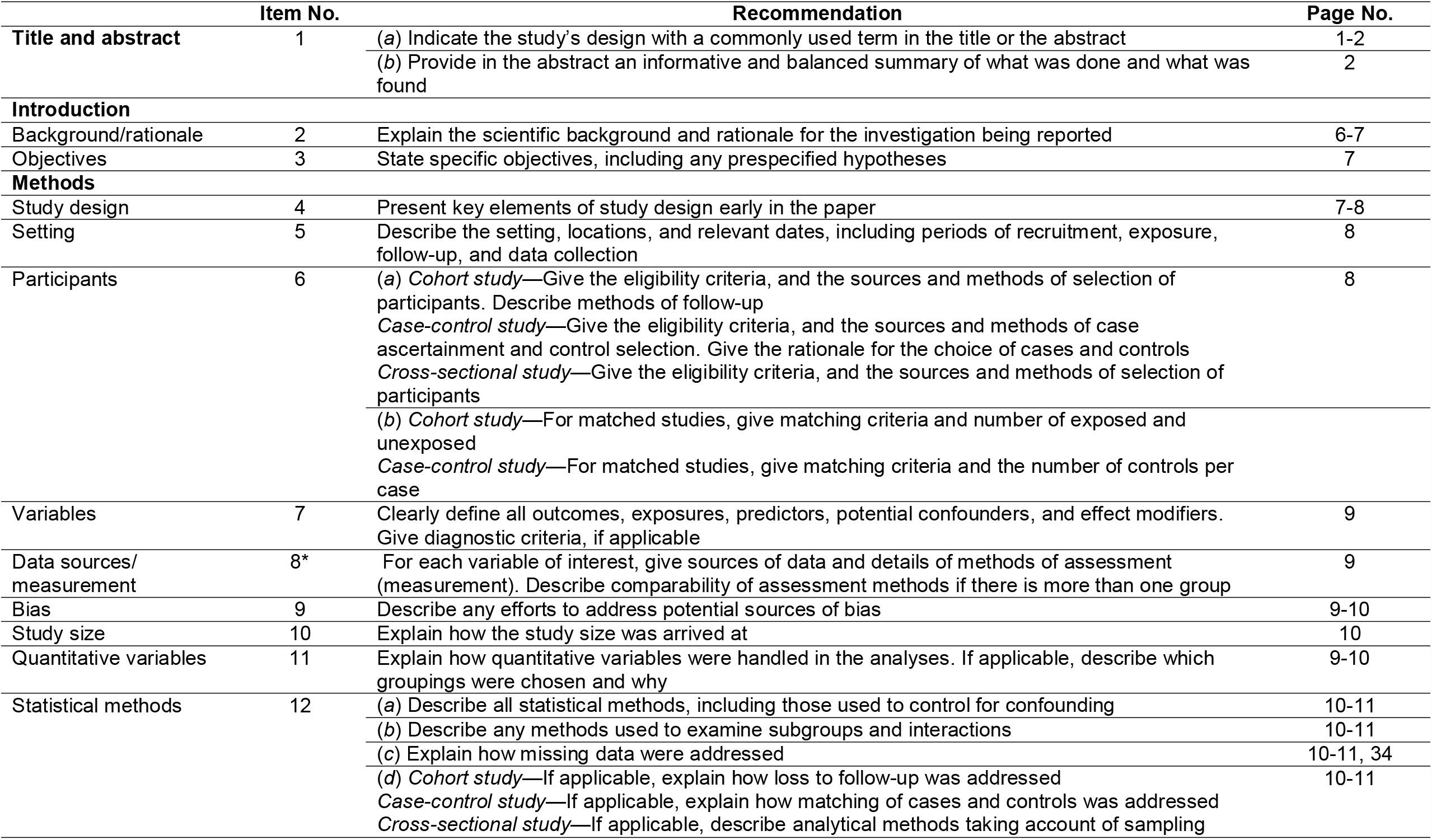
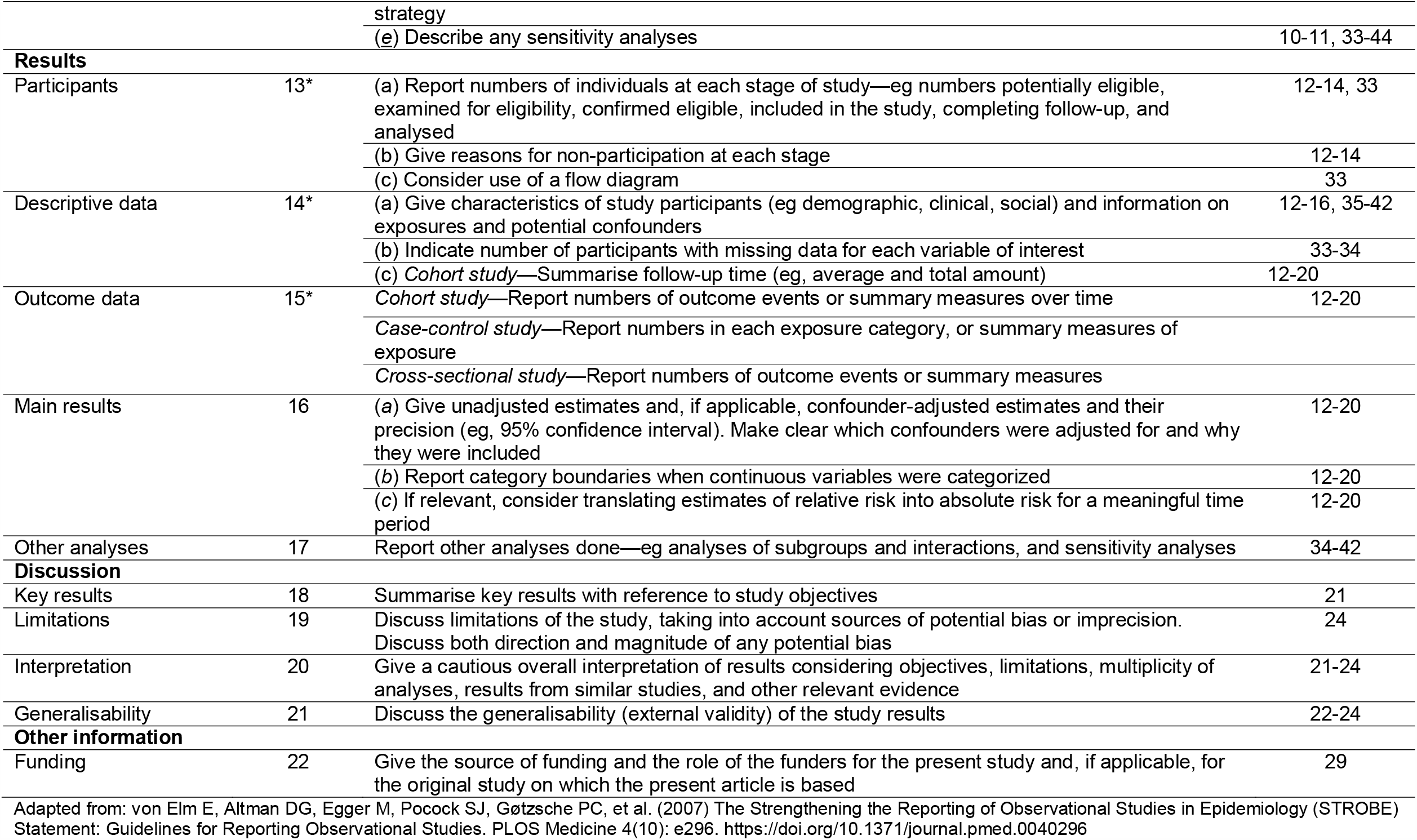

|  | Item No. | Recommendation | Page No. |
| --- | --- | --- | --- |
| Title and abstract | 1 | (a) Indicate the study's design with a commonly used term in the title or the abstract | 1-2 |
|  |  | (b) Provide in the abstract an informative and balanced summary of what was done and what was found | 2 |
| Introduction |  |  |  |
| Background/rationale | 2 | Explain the scientific background and rationale for the investigation being reported | 6-7 |
| Objectives | 3 | State specific objectives, including any prespecified hypotheses | 7 |
| Methods |  |  |  |
| Study design | 4 | Present key elements of study design early in the paper | 7-8 |
| Setting | 5 | Describe the setting, locations, and relevant dates, including periods of recruitment, exposure, follow-up, and data collection | 8 |
| Participants | 6 | (a) Cohort study—Give the eligibility criteria, and the sources and methods of selection of participants. Describe methods of follow-up | 8 |
|  |  | Case-control study—Give the eligibility criteria, and the sources and methods of case ascertainment and control selection. Give the rationale for the choice of cases and controls |  |
|  |  | Cross-sectional study—Give the eligibility criteria, and the sources and methods of selection of participants |  |
|  |  | (b) Cohort study—For matched studies, give matching criteria and number of exposed and unexposed |  |
|  |  | Case-control study—For matched studies, give matching criteria and the number of controls per case |  |
| Variables | 7 | Clearly define all outcomes, exposures, predictors, potential confounders, and effect modifiers. Give diagnostic criteria, if applicable | 9 |
| Data sources/measurement | 8* | For each variable of interest, give sources of data and details of methods of assessment (measurement). Describe comparability of assessment methods if there is more than one group | 9 |
| Bias | 9 | Describe any efforts to address potential sources of bias | 9-10 |
| Study size | 10 | Explain how the study size was arrived at | 10 |
| Quantitative variables | 11 | Explain how quantitative variables were handled in the analyses. If applicable, describe which groupings were chosen and why | 9-10 |
| Statistical methods | 12 | (a) Describe all statistical methods, including those used to control for confounding | 10-11 |
|  |  | (b) Describe any methods used to examine subgroups and interactions | 10-11 |
|  |  | (c) Explain how missing data were addressed | 10-11, 34 |
|  |  | (d) Cohort study—If applicable, explain how loss to follow-up was addressed | 10-11 |
|  |  | Case-control study—If applicable, explain how matching of cases and controls was addressed |  |
|  |  | Cross-sectional study—If applicable, describe analytical methods taking account of sampling strategy |  |
|  |  | (e) Describe any sensitivity analyses | 10-11, 33-44 |
| **Results** |  |  |  |
| Participants | 13* | (a) Report numbers of individuals at each stage of study—eg numbers potentially eligible, examined for eligibility, confirmed eligible, included in the study, completing follow-up, and analysed | 12-14, 33 |
|  |  | (b) Give reasons for non-participation at each stage | 12-14 |
|  |  | (c) Consider use of a flow diagram | 33 |
| Descriptive data | 14* | (a) Give characteristics of study participants (eg demographic, clinical, social) and information on exposures and potential confounders | 12-16, 35-42 |
|  |  | (b) Indicate number of participants with missing data for each variable of interest | 33-34 |
|  |  | (c) *Cohort study*—Summarise follow-up time (eg, average and total amount) | 12-20 |
| Outcome data | 15* | *Cohort study*—Report numbers of outcome events or summary measures over time | 12-20 |
|  |  | *Case-control study*—Report numbers in each exposure category, or summary measures of exposure |  |
|  |  | *Cross-sectional study*—Report numbers of outcome events or summary measures |  |
| Main results | 16 | (a) Give unadjusted estimates and, if applicable, confounder-adjusted estimates and their precision (eg, 95% confidence interval). Make clear which confounders were adjusted for and why they were included | 12-20 |
|  |  | (b) Report category boundaries when continuous variables were categorized | 12-20 |
|  |  | (c) If relevant, consider translating estimates of relative risk into absolute risk for a meaningful time period | 12-20 |
| Other analyses | 17 | Report other analyses done—eg analyses of subgroups and interactions, and sensitivity analyses | 34-42 |
| **Discussion** |  |  |  |
| Key results | 18 | Summarise key results with reference to study objectives | 21 |
| Limitations | 19 | Discuss limitations of the study, taking into account sources of potential bias or imprecision. Discuss both direction and magnitude of any potential bias | 24 |
| Interpretation | 20 | Give a cautious overall interpretation of results considering objectives, limitations, multiplicity of analyses, results from similar studies, and other relevant evidence | 21-24 |
| Generalisability | 21 | Discuss the generalisability (external validity) of the study results | 22-24 |
| **Other information** |  |  |  |
| Funding | 22 | Give the source of funding and the role of the funders for the present study and, if applicable, for the original study on which the present article is based | 29 |

Adapted from: von Elm E, Altman DG, Egger M, Pocock SJ, Gøtzsche PC, et al. (2007) The Strengthening the Reporting of Observational Studies in Epidemiology (STROBE) Statement: Guidelines for Reporting Observational Studies. PLOS Medicine 4(10): e296. https://doi.org/10.1371/journal.pmed.0040296

## APPENDIX 2: ETHICAL APPROVAL

Ethical approval for this study was granted by the following ethics committees.

| Committee | Reference |
| --- | --- |
| *United Kingdom* |  |
| University College London Research Ethics Committee | 17123/001 |
| *Malawi* |  |
| College of Medicine Research and Ethics Committee | P.01/20/2909 |
| *Zimbabwe* |  |
| Medical Research Council of Zimbabwe | MRCZ/A/2570 |
| Joint Research Ethics Committee for the University of Zimbabwe,<br>College of Health Sciences and Parirenyatwa Group of Hospitals | JREC/327/19 |
| Biomedical Research and Training Institute Institutional Review Board | AP155/2020 |
| Sally Mugabe (Harare) Central Hospital Ethics Committee | 071119/64 |

## APPENDIX 3: FLOW DIAGRAMS OF RECORD INCLUSION

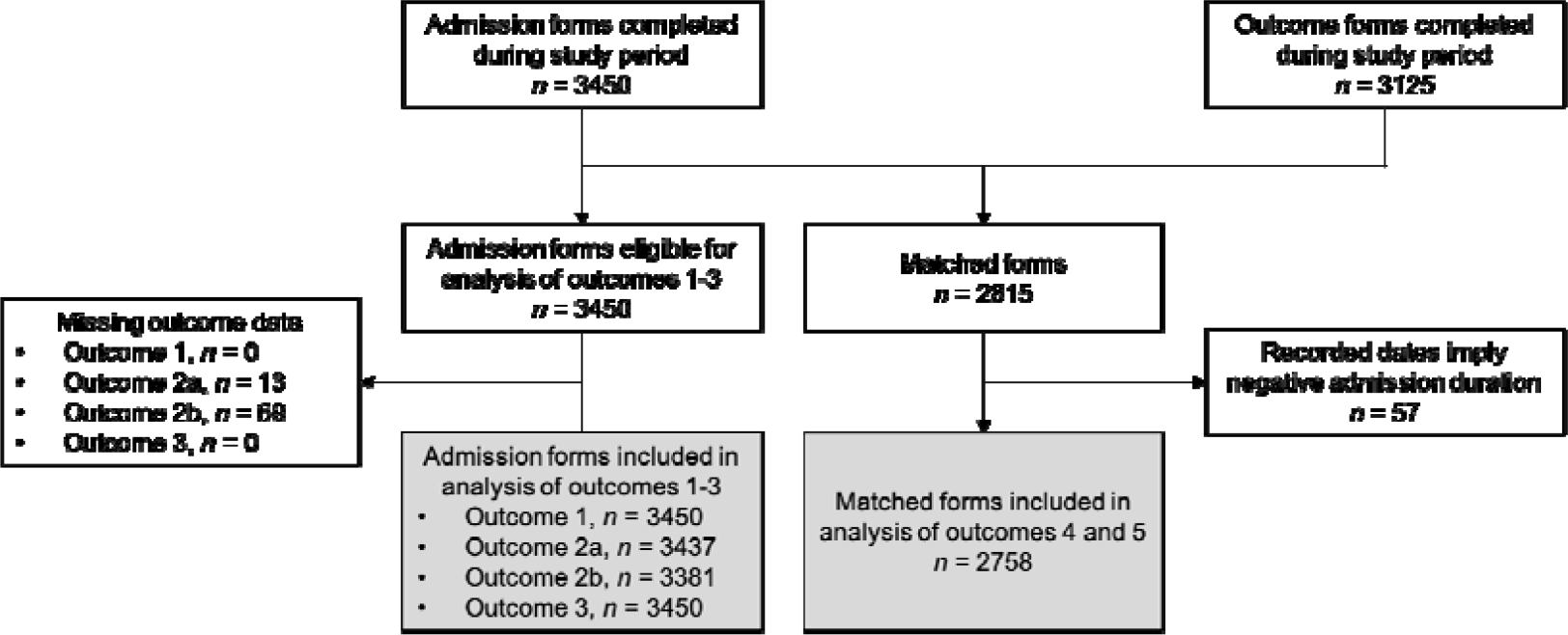

**Flow diagram of record inclusion for analysis of data at Sally Mugabe Central Hospital, Zimbabwe**

- Outcome 1: number of admissions
- Outcome 2a: gestational age
- Outcome 2b: birth weight
- Outcome 3: source of admission
- Outcome 4: prevalence of neonatal encephalopathy
- Outcome 5: overall mortality rate

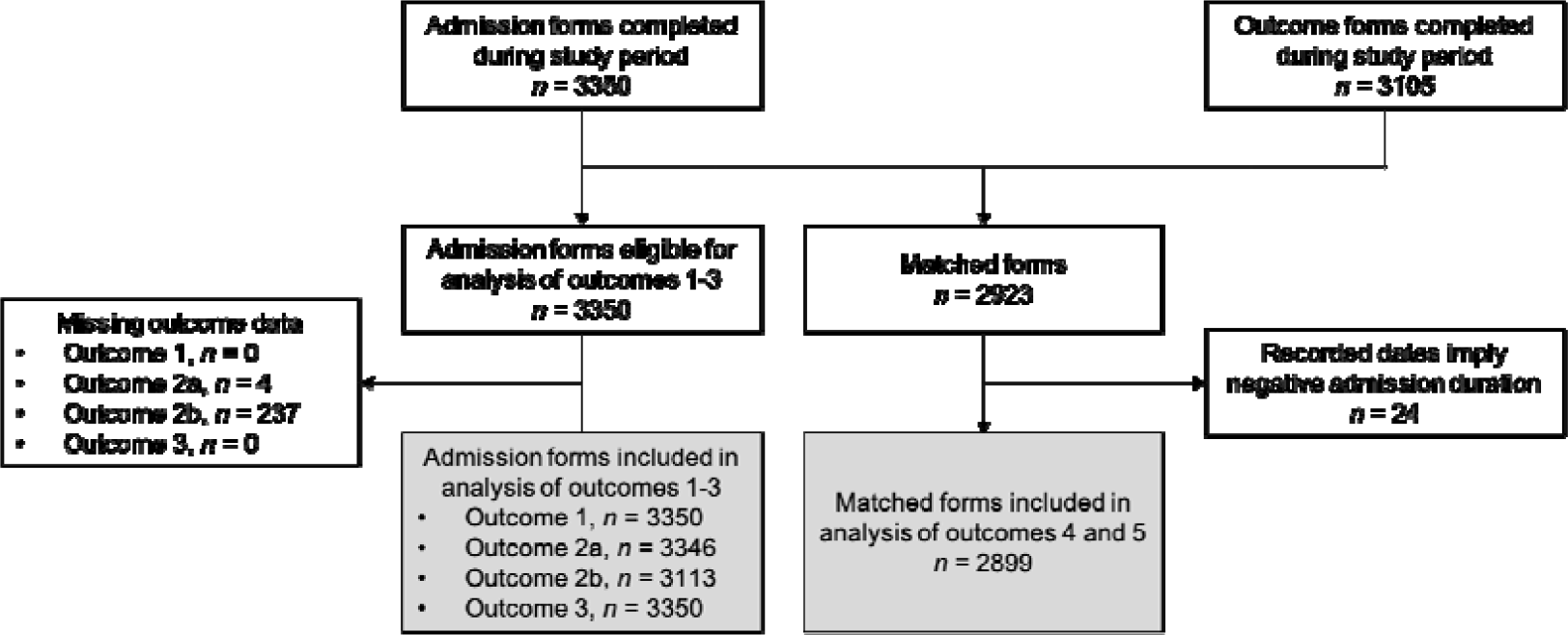

**Flow diagram of record inclusion for analysis of data at Kamuzu Central Hospital, Malawi**

- Outcome 1: number of admissions
- Outcome 2a: gestational age
- Outcome 2b: birth weight
- Outcome 3: source of admission
- Outcome 4: prevalence of neonatal encephalopathy
- Outcome 5: overall mortality rate

## APPENDIX 4: MISSING DATA

The table below shows the number of participants with missing data for each outcome and the number of participants remaining for each analysis after pairwise deletion of missing values.

| Characteristics | *n* missing (%) |  | *n* remaining* |  |
| --- | --- | --- | --- | --- |
|  | SMCH | KCH | SMCH | KCH |
| Gestational age | 13 (0.4) | 4 (0.1) | 3437 (99.6) | 3346 (99.9) |
| Birth weight | 69 (2.0) | 237 (7.1) | 3381 (98.0) | 3113 (92.9) |
| Source of admission | 0 (0.0) | 0 (0.0) | 3450 (100.0) | 3350 (100.0) |
| Neonatal encephalopathy | 0 (0.0) | 0 (0.0) | 2758 (100.0)† | 2899 (100.0)† |
| Death | 0 (0.0) | 0 (0.0) | 2758 (100.0)† | 2899 (100.0)† |

- \* Remaining for analysis after pairwise deletion. - † Only matched admission and outcome forms considered for analysis of neonatal encephalopathy and death. - SMCH: Sally Mugabe Central Hospital; KCH: Kamuzu Central Hospital, Malawi

## APPENDIX 5: FURTHER REGRESSION ANALYSIS RESULTS

### Outcome 1: Admissions to the neonatal unit

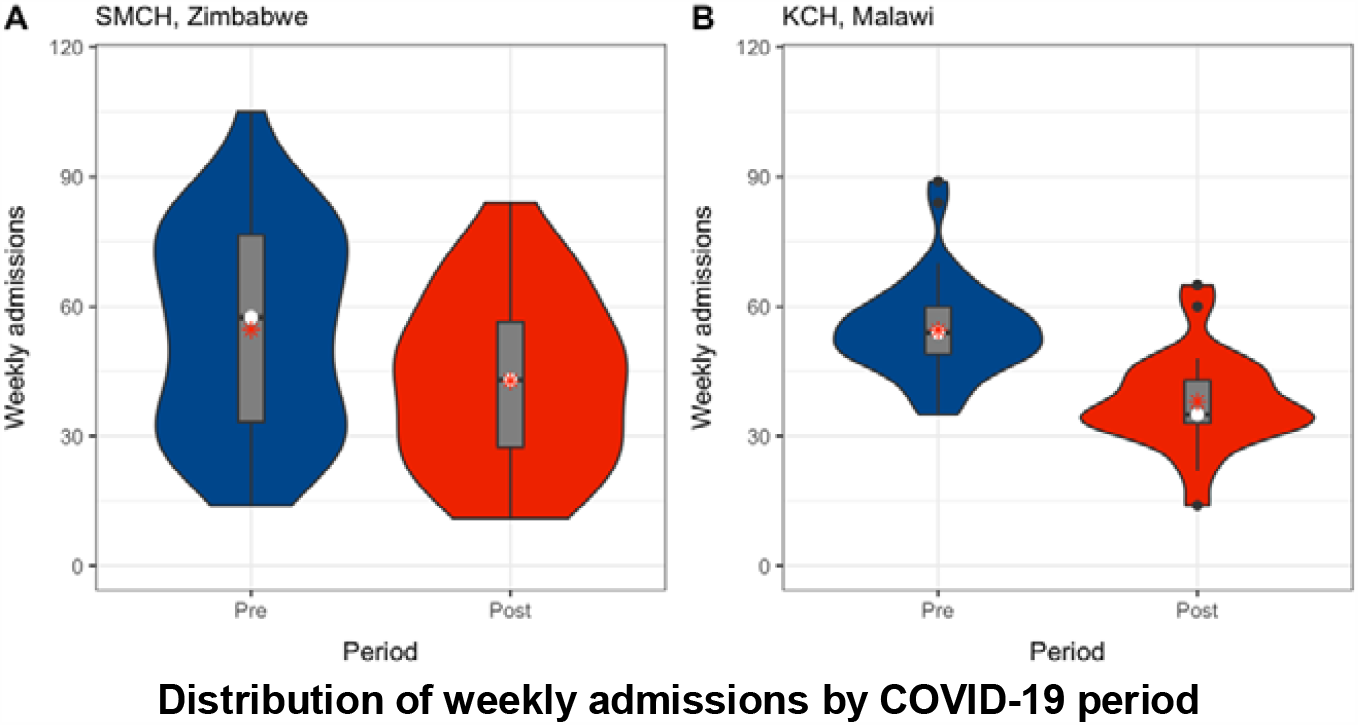

**SMCH model 1: Level change model, adjusted for doctors’ strike period**

SMCH model 1: Level change model, adjusted for doctors' strike period
|  | Coef | SE | Exp | 95% CI | p-value |
| --- | --- | --- | --- | --- | --- |
| Intercept | 4.43 | 0.08 | 84.31 | 71.89 – 98.86 | < 0.001 |
| Post-COVID-19 period, yes | -0.19 | 0.17 | 0.83 | 0.60 – 1.14 | 0.25 |
| Study time elapsed, weeks | -0.009 | 0.004 | 0.99 | 0.98 – 1.00 | 0.012 |
| Doctors' strike period, yes | -0.68 | 0.11 | 0.51 | 0.41 – 0.63 | < 0.001 |

**SMCH model 2: Level change model, additionally adjusted for nurses’ strike period**

SMCH model 2: Level change model, additionally adjusted for nurses' strike period
|  | Coef | SE | Exp | 95% CI | p-value |
| --- | --- | --- | --- | --- | --- |
| Intercept | 4.37 | 0.07 | 79.20 | 68.81 – 91.16 | < 0.001 |
| Post- COVID-19 period, yes | -0.11 | 0.14 | 0.90 | 0.69 – 1.17 | 0.90 |
| Study time elapsed, weeks | -0.005 | 0.003 | 1.00 | 0.99 – 1.00 | 0.10 |
| Doctors' strike period, yes | -0.70 | 0.09 | 0.50 | 0.41 – 0.60 | < 0.001 |
| Nurses' strike period, yes | -0.65 | 0.14 | 0.52 | 0.40 – 0.68 | < 0.001 |

**KCH model: Level change model, unadjusted**

|  | Coef | SE | Exp | 95% CI | p-value |
| --- | --- | --- | --- | --- | --- |
| Intercept | 3.88 | 0.06 | 48.42 | 43.03 – 54.49 | < 0.001 |
| Post- COVID-19 period, yes | -0.55 | 0.10 | 0.58 | 0.48 – 0.70 | < 0.001 |
| Study time elapsed, weeks | 0.005 | 0.002 | 1.01 | 1.00 – 1.01 | 0.019 |

### Outcome 2: Gestational age at birth and birth weight

#### Gestational age at birth

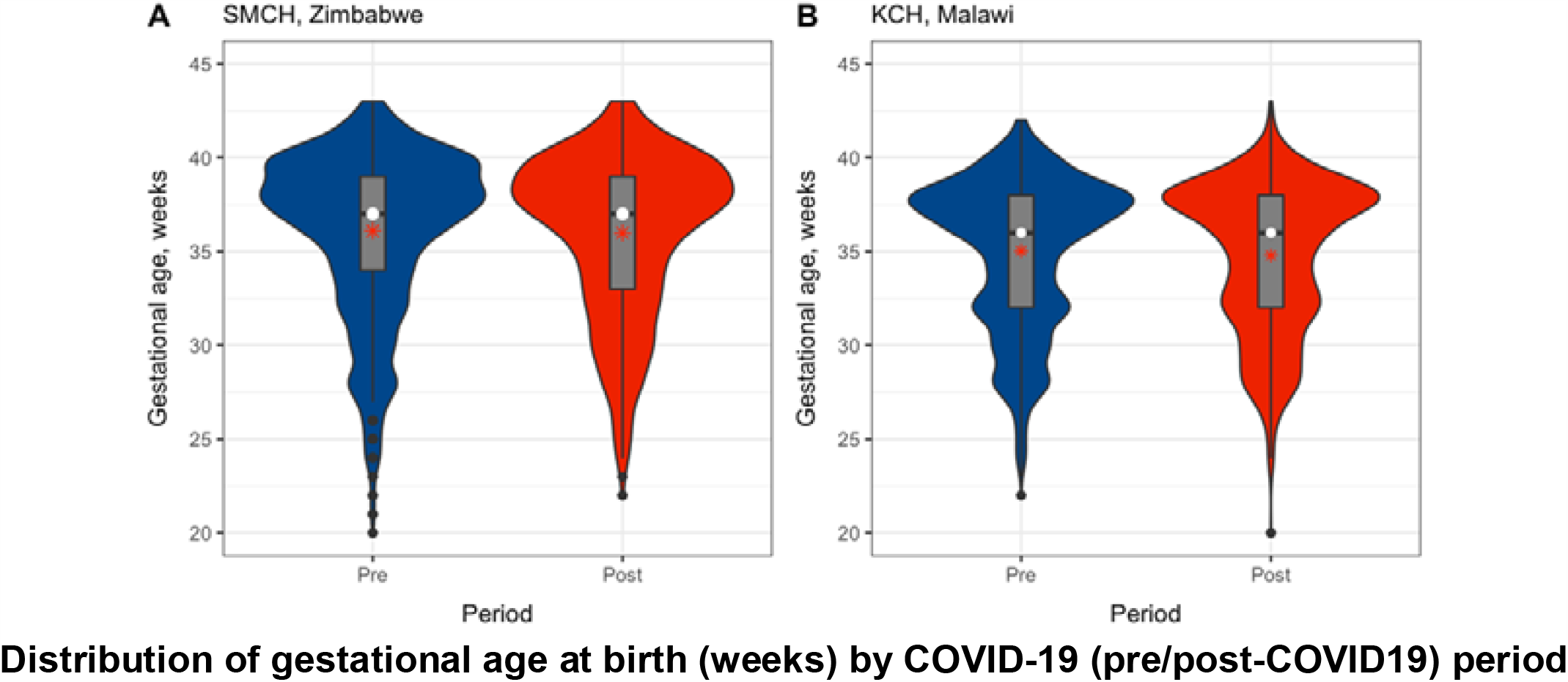

**SMCH model: Level change model, adjusted for doctors’ strike period**

**SMCH model: Level change model, adjusted for doctors' strike period**
|  | Coef | SE | 95% CI | p-value |
| --- | --- | --- | --- | --- |
| Intercept | 36.25 | 0.15 | 35.96 – 36.54 | < 0.001 |
| Post-COVID-19 period, yes | 0.04 | 0.29 | -0.53 – 0.61 | 0.89 |
| Study time elapsed, weeks | -0.006 | 0.006 | -0.02 – 0.007 | 0.37 |
| Doctors' strike period, yes | -0.17 | 0.20 | -0.57 – 0.23 | 0.41 |

**KCH model: Level change model, unadjusted**

|  | Coef | SE | 95% CI | p-value |
| --- | --- | --- | --- | --- |
| Intercept | 34.42 | 0.15 | 34.12 – 34.72 | < 0.001 |
| Post-COVID-19 period, yes | -1.14 | 0.25 | -1.62 – -0.65 | < 0.001 |
| Study time elapsed, weeks | 0.03 | 0.006 | 0.02 – 0.04 | < 0.001 |

#### Birth weight

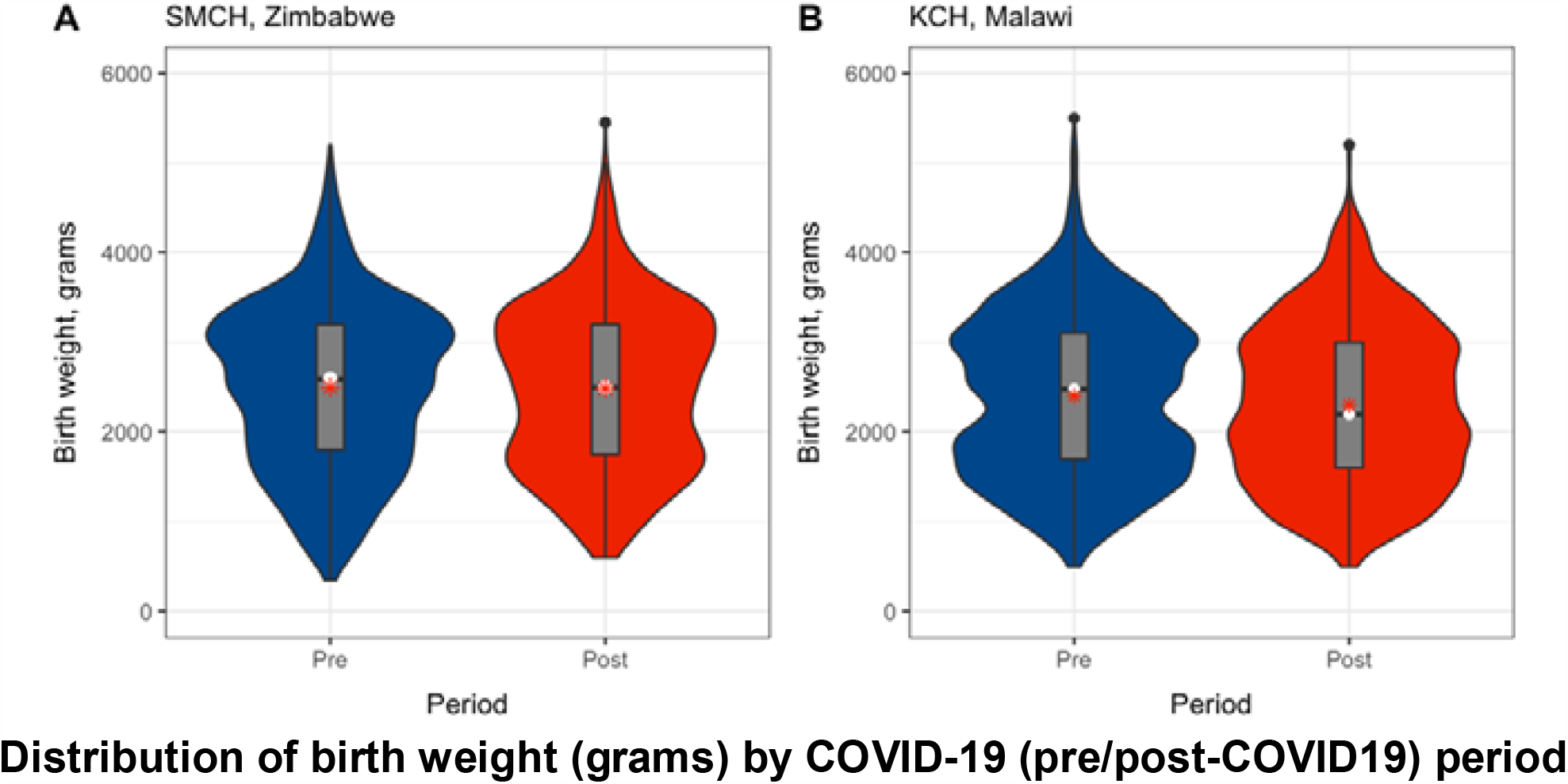

**SMCH model: Level change model, adjusted for doctors’ strike period**

SMCH model: Level change model, adjusted for doctors' strike period
|  | Coef | SE | 95% CI | p-value |
| --- | --- | --- | --- | --- |
| Intercept | 2530.0 | 31.5 | 2468.0 – 2591.4 | < 0.001 |
| Post- COVID-19 period, yes | -7.2 | 61.1 | -127.1 – 112.6 | 0.91 |
| Study time elapsed, weeks | -0.7 | 1.3 | -3.3 – 2.0 | 0.62 |
| Doctors' strike period, yes | -58.1 | 42.9 | -142.1 – 25.9 | 0.18 |

**KCH model: Level change model, unadjusted**

|  | Coef | SE | 95% CI | p-value |
| --- | --- | --- | --- | --- |
| Intercept | 2269.0 | 36.0 | 2198.4 – 2339.6 | < 0.001 |
| Post- COVID-19 period, yes | -299.9 | 57.3 | -412.3 – -187.5 | < 0.001 |
| Study time elapsed, weeks | 5.9 | 1.4 | 3.2 – 8.6 | < 0.001 |

### Outcome 3: Source of admission referral

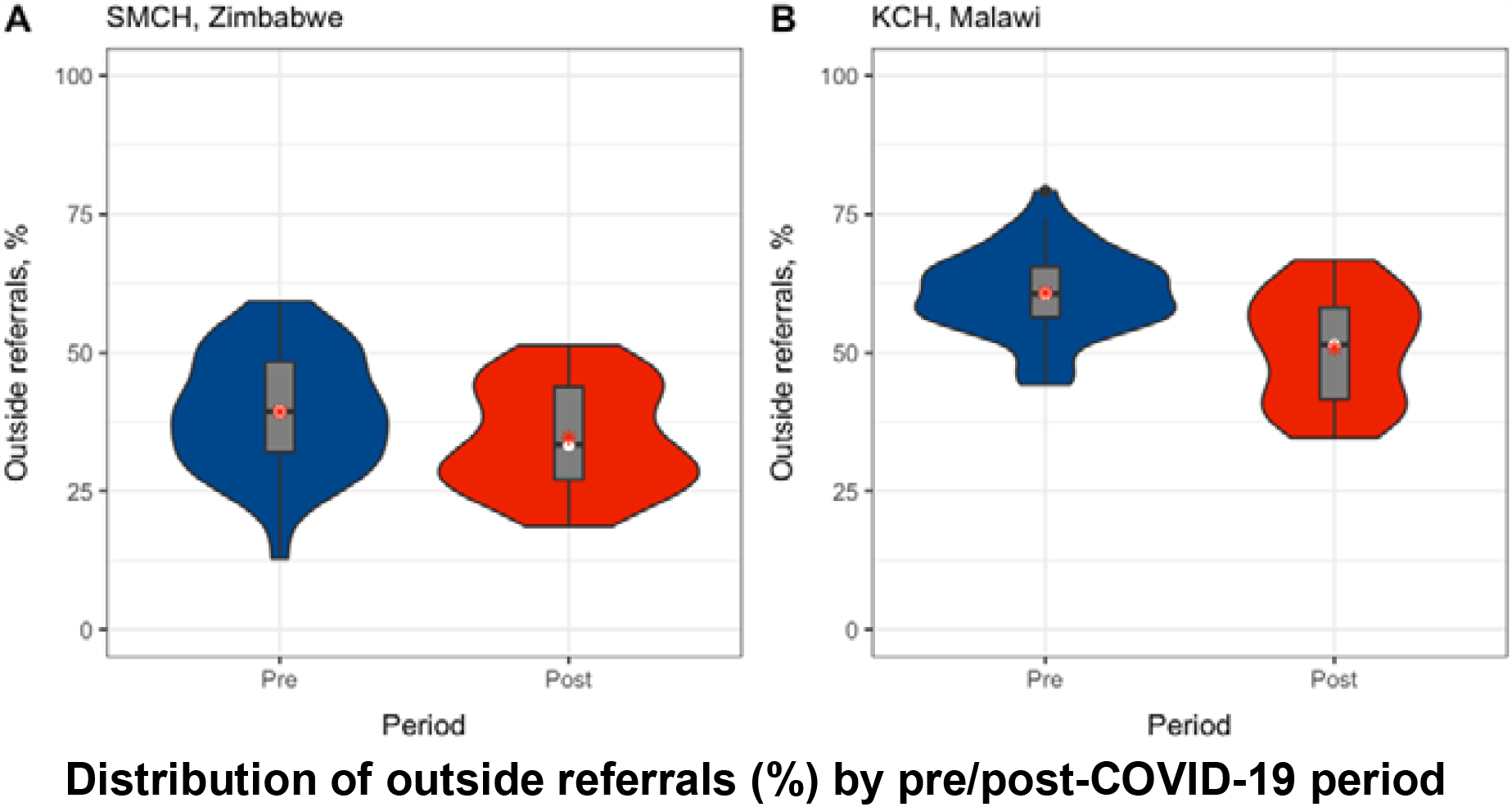

**SMCH model: Level change model, adjusted for doctors’ strike period**

SMCH model: Level change model, adjusted for doctors' strike period
|  | Coef | SE | Exp | 95% CI | p-value |
| --- | --- | --- | --- | --- | --- |
| Intercept | -1.15 | 0.06 | 0.32 | 0.28 – 0.36 | < 0.001 |
| Post- COVID-19 period, yes | -0.02 | 0.11 | 0.98 | 0.79 – 1.23 | 0.88 |
| Study time elapsed, weeks | 0.001 | 0.003 | 1.00 | 1.00 – 1.01 | 0.55 |
| Doctors' strike period, yes | 0.33 | 0.07 | 1.39 | 1.20 – 1.60 | < 0.001 |

**KCH model: Level change model, unadjusted**

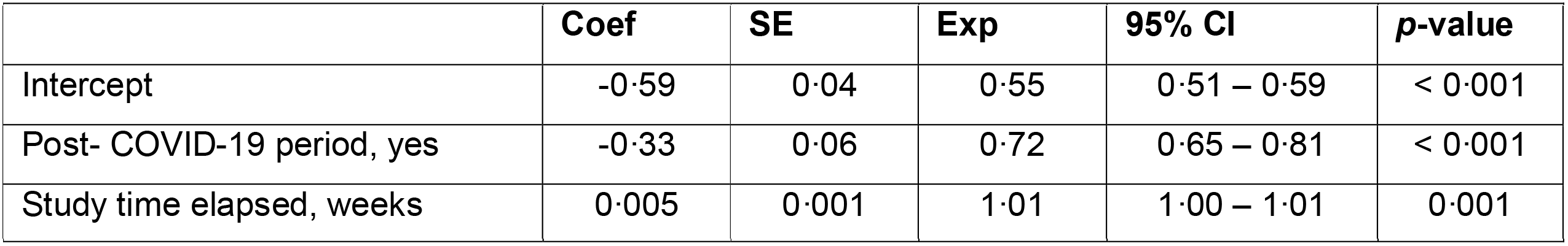

|  | Coef | SE | Exp | 95% CI | p-value |
| --- | --- | --- | --- | --- | --- |
| Intercept | -0.59 | 0.04 | 0.55 | 0.51 – 0.59 | < 0.001 |
| Post- COVID-19 period, yes | -0.33 | 0.06 | 0.72 | 0.65 – 0.81 | < 0.001 |
| Study time elapsed, weeks | 0.005 | 0.001 | 1.01 | 1.00 – 1.01 | 0.001 |

### Outcome 4: Prevalence of neonatal encephalopathy

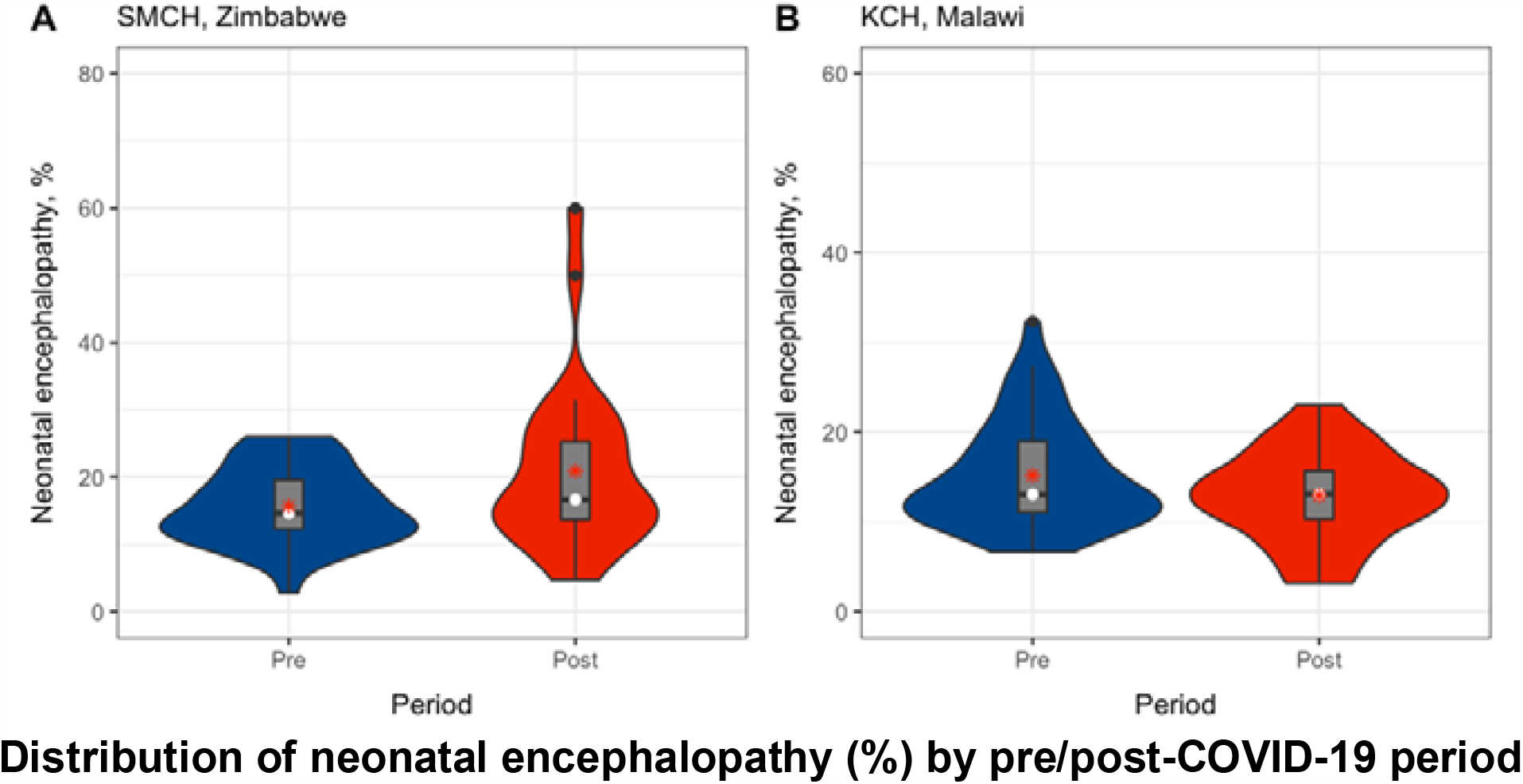

**SMCH model: Level change model, adjusted for doctors’ strike period**

SMCH model: Level change model, adjusted for doctors' strike period
|  | Coef | SE | Exp | 95% CI | p-value |
| --- | --- | --- | --- | --- | --- |
| Intercept | -1.93 | 0.10 | 0.15 | 0.12 – 0.18 | < 0.001 |
| Post- COVID-19 period, yes | 0.08 | 0.18 | 1.08 | 0.76 – 1.55 | 0.67 |
| Study time elapsed, weeks | 0.004 | 0.004 | 1.00 | 1.00 – 1.01 | 0.27 |
| Doctors' strike period, yes | -0.02 | 0.13 | 0.98 | 0.76 – 1.26 | 0.87 |

**KCH model: Level change model, unadjusted**

|  | Coef | SE | Exp | 95% CI | p-value |
| --- | --- | --- | --- | --- | --- |
| Intercept | -1.66 | 0.09 | 0.19 | 0.16 – 0.23 | < 0.001 |
| Post- COVID-19 period, yes | 0.27 | 0.16 | 1.30 | 0.95 – 1.80 | 0.11 |
| Study time elapsed, weeks | -0.01 | 0.004 | 0.99 | 0.98 – 1.00 | 0.001 |

### Outcome 5: Overall mortality

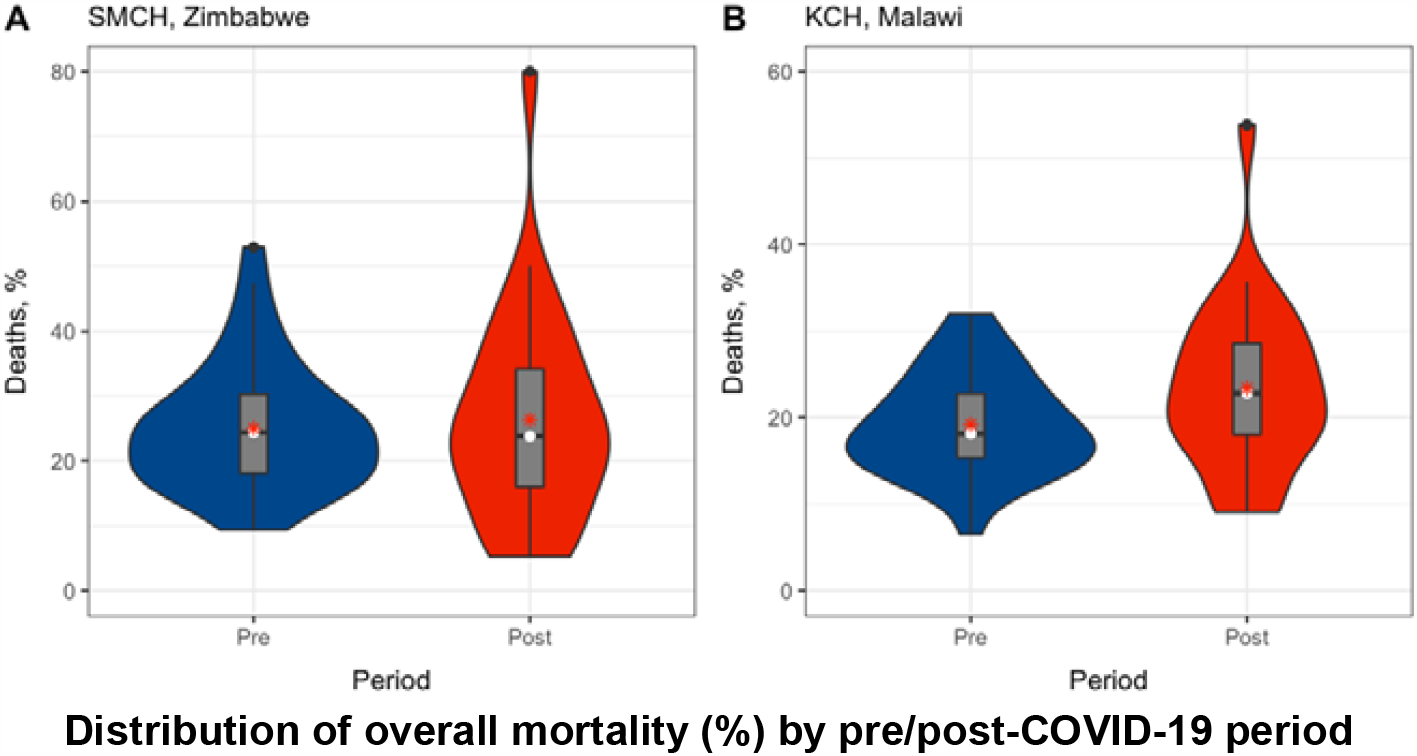

**SMCH model 1: Level change model, adjusted for doctors’ strike period**

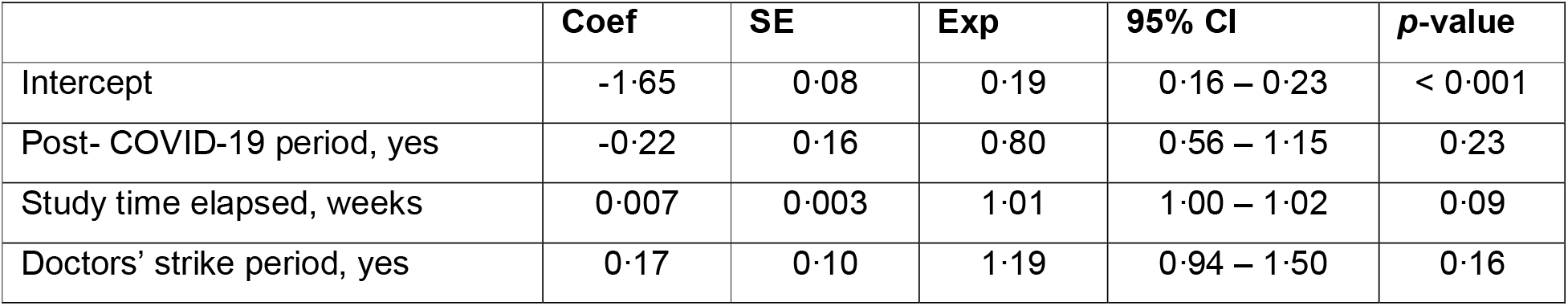

**SMCH model 2: Level change model, additionally adjusted for nurses’ strike period**

SMCH model 2: Level change model, additionally adjusted for nurses' strike period
|  | Coef | SE | Exp | 95% CI | p-value |
| --- | --- | --- | --- | --- | --- |
| Intercept | -1.60 | 0.09 | 0.20 | 0.17 – 0.24 | < 0.001 |
| Post- COVID-19 period, yes | -0.33 | 0.18 | 0.72 | 0.51 – 1.03 | 0.07 |
| Study time elapsed, weeks | 0.004 | 0.004 | 1.00 | 1.00 – 1.01 | 0.30 |
| Doctors' strike period, yes | 0.19 | 0.11 | 1.21 | 0.98 – 1.50 | 0.08 |
| Nurses' strike period, yes | 0.60 | 0.17 | 1.82 | 1.30 – 2.55 | 0.001 |

**KCH model 1: Level change model, unadjusted**

| Mal – deaths (unadjusted) | Coef | SE | Exp | 95% CI | p-value |
| --- | --- | --- | --- | --- | --- |
| Intercept | -1.56 | 0.09 | 0.21 | 0.18 – 0.25 | < 0.001 |
| Post- COVID-19 period, yes | 0.27 | 0.14 | 1.31 | 0.98 – 1.73 | 0.07 |
| Study time elapsed, weeks | -0.004 | 0.003 | 1.00 | 0.99 – 1.00 | 0.27 |

**KCH model 2: Slope change model, unadjusted**

| **Mal – deaths (sensitivity)** | **Coef** | **SE** | **Exp** | **95% CI** | **p-value** |
| --- | --- | --- | --- | --- | --- |
| Intercept | -1.55 | 0.09 | 0.21 | 0.18 – 0.25 | < 0.001 |
| Study time elapsed, weeks | -0.004 | 0.003 | 1.00 | 0.99 – 1.00 | 0.25 |
| Time since first COVID-19 case,<br>weeks * post- COVID-19 period,<br>yes | 0.02 | 0.009 | 1.02 | 1.00 – 1.04 | 0.04 |

## APPENDIX 6: ADDITIONAL ANALYSES

### Mode of delivery of admitted neonates

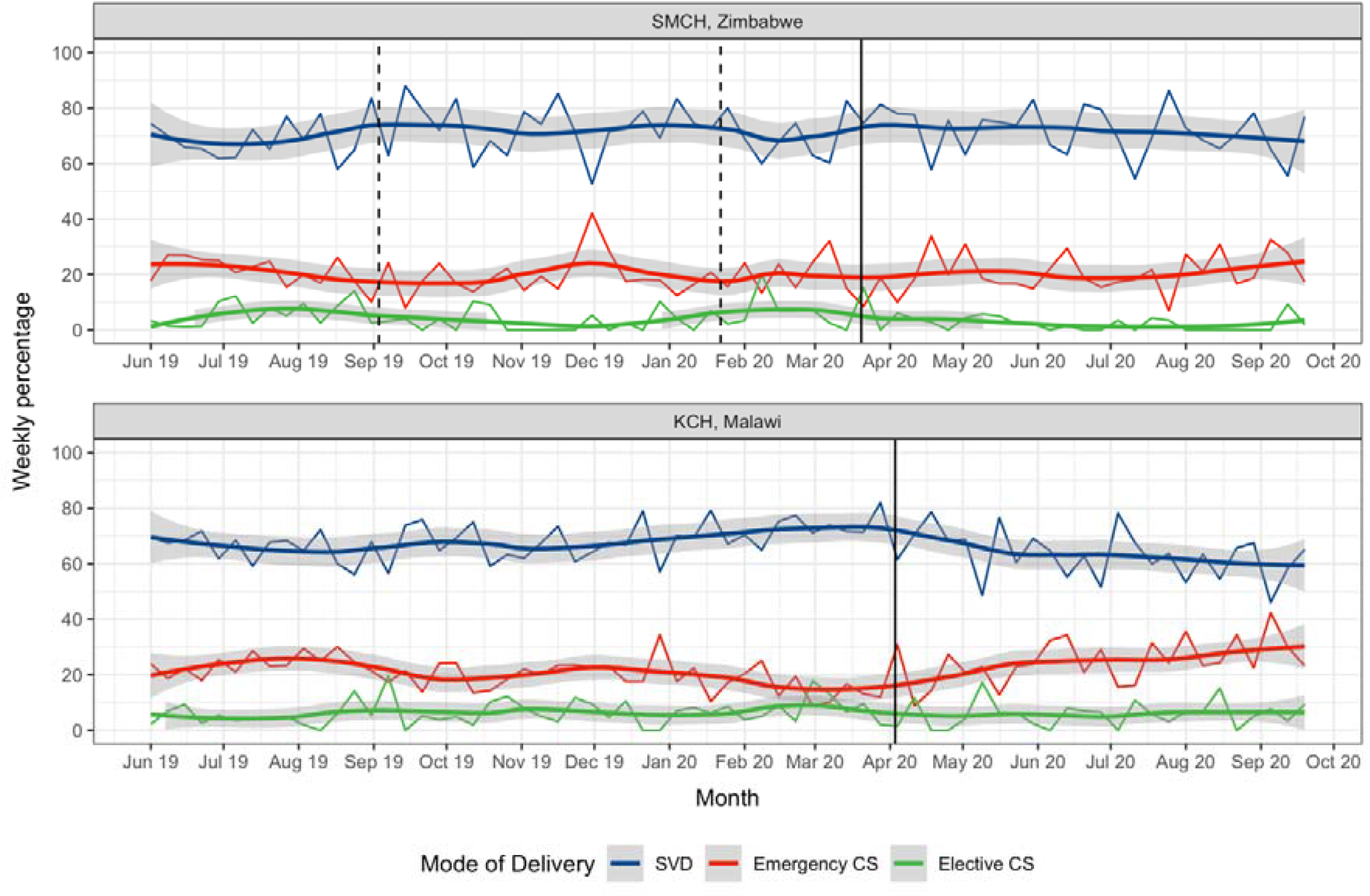

**Trend in mode of delivery of admitted neonates per week**

- Only SVD, emergency CS and elective CS displayed here to avoid overplotting.
- Smoothed line: local regression (LOESS) model; shaded region: 95% confidence interval.
- Solid vertical line: first confirmed case of COVID-19 in each country.
- Period between dashed vertical lines: industrial action by doctors in Zimbabwe.
- Counts based on all admission forms completed, irrespective of match status.
- *SMCH: Sally Mugabe Central Hospital; KCH: Kamuzu Central Hospital; SVD: spontaneous vaginal delivery; CS: caesarean section*

